# Multiple pathways to metabolic syndrome identified by hierarchical clustering of Japanese longitudinal health checkup data

**DOI:** 10.64898/2026.03.06.26347135

**Authors:** Satomi Shimmura, Makito Oku, Yoshiki Nagata, Takashi Yamagami, Shiho Fujisaka, Tsutomu Wada, Toshiyasu Sasaoka, Yasuhiro Onogi, Hisae Honoki, Tomonobu Kado, Ayumi Nishimura, Muhammad Bilal, Michikazu Sekine, Kazuyuki Tobe

## Abstract

To identify pre-onset pathways of metabolic syndrome (MetS), we retrospectively analyzed Japanese health checkup data from 296 male MetS cases and 296 non-MetS controls matched by initial status (aged 49–64 years). Hierarchical clustering was applied to four MetS components—abdominal obesity (OB), hypertension (HT), dyslipidemia (DLP), and hyperglycemia (HG)—using longitudinal status from the 3 years (years −3 to −1) before onset (year 0). Six clusters were identified, showing shifts from early (year −3 to −1) to late (year −1 to 0) patterns. Cluster 1 (n = 97, 32.8%): early OB and HT; followed by DLP. Cluster 2 (n = 36, 12.2%): early OB and DLP; followed by HT. Cluster 3 (n = 48, 16.2%): early OB; followed by HT and HG. Cluster 4 (n = 44, 14.9%): early HT and DLP; followed by OB. Cluster 5 (n = 46, 15.5%): early HT; followed by OB, DLP, and HG. Cluster 6 (n = 25, 8.4%): early HT, DLP, and HG; followed by OB. Controls showed no clear changes. The waist circumference and triglyceride-glucose related indices were significantly higher at year 0 in the MetS group. These findings provide insight into the heterogeneous pathways leading to MetS. This clustering approach offers a flexible framework for capturing population-specific patterns in MetS development and may allow more targeted early intervention strategies.

## Introduction

Metabolic syndrome (MetS) is a constellation of interrelated abnormalities generally associated with insulin resistance, a condition closely linked to abdominal obesity (OB). These abnormalities—which are referred to as MetS components in this paper, including OB, hypertension (HT), dyslipidemia (DLP), and hyperglycemia (HG)—synergistically contribute to a heightened risk of cardiovascular disease and type 2 diabetes [1–3]. Early prevention of MetS is essential and understanding the progression dynamics prior to MetS onset is crucial for effective intervention.

Clustering methods have been applied to identify pathways to disease, contributing to mechanistic insights across various conditions [4–10]. In studies of MetS, clustering approaches have provided valuable insights into clinical outcomes [7–10]. However, early-stage trajectories leading to MetS remain underexplored. Few studies have tracked individuals over time to capture the temporal evolution of MetS components.

On the other hand, previous studies have attempted to analyze the natural history of MetS using Markov models [11,12], which simulate disease progression by modeling transitions between discrete health states based on transition probabilities [13–16]. While Markov models offer valuable insights into disease progression, they are not as powerful as clustering methods to distinguish multiple pathways.

We hypothesized that multiple pathways exist prior to MetS onset, and that machine learning–based approaches could help elucidate these trajectories. Based on these expectations, we applied clustering analysis to Japanese longitudinal health checkup data on MetS components preceding diagnosis. By comparing these trajectories with those of non-MetS controls who had similar clinical profiles to those of MetS patients three years before diagnosis, we aimed to characterize the temporal patterns of MetS components within each cluster and compare these patterns across clusters.

## Methods

The following abbreviations are used throughout this study: metabolic syndrome (MetS), abdominal obesity (OB), hypertension (HT), dyslipidemia (DLP), hyperglycemia (HG), waist circumference (WC), body mass index (BMI), systolic blood pressure (SBP), diastolic blood pressure (DBP), triglycerides (TG), high-density lipoprotein cholesterol (HDL-C), plasma glucose (PG), glycated hemoglobin (HbA1c), triglyceride-glucose index × waist circumference (TyG-WC), and triglyceride-glucose index × body mass index (TyG-BMI).

### Definition and diagnostic criteria for MetS

MetS was diagnosed as the following, based on the requirement of OB, with reference to the Japanese criteria [17]. The presence of MetS was determined for each participant at each time point. The year of onset was defined as year 0. Diagnosis required OB plus at least two of the following components: HT, DLP, or HG. OB was defined as a WC ≥ 85 cm for males or ≥ 90 cm for females. The additional components were defined as follows: HT: SBP ≥ 130 mmHg, DBP ≥ 85 mmHg, or current use of antihypertensive medication. DLP: TG ≥ 150 mg/dL, HDL-C < 40 mg/dL, or current use of lipid-lowering medication. HG: Fasting PG ≥ 110 mg/dL, HbA1c ≥ 6.0%, current or past use of antidiabetic medication, or a clinical diagnosis of diabetes. Diabetes was defined with reference to global standards [18] as fasting PG ≥ 126 mg/dL or HbA1c ≥ 6.5%, current or past use of antidiabetic medication, or a clinical diagnosis of diabetes, either ongoing or previously established.

### Primary variables used in the analysis

We analyzed 11 numerical variables of health checkup data: age, WC, BMI, SBP, DBP, TG, HDL-C, PG, HbA1c, TyG-WC, and TyG-BMI.

We also analyzed several categorical variables. Information on smoking status, alcohol consumption, intention to improve lifestyle habits, and willingness to receive specific health guidance was obtained from a standardized self-administered questionnaire.

Responses were converted into binary variables and used to define derived variables for multivariable analyses. Details of questionnaire coding and variable definitions are provided in S1–S3 Tables.

All measurements and questionnaire assessments were conducted using identical standardized protocols defined in the Standard Program for Specific Health Checkups and Guidance [19,20], ensuring comparability across all participants.

### Encoding of MetS components

Each MetS component was categorized into three levels (0, normal, 1, intermediate, and 2, MetS criteria) to harmonize variables with different measurement units, using thresholds often applied for each MetS component. Specific thresholds for each level are provided in S4 Table. These three-level encoded variables were used in both the clustering analysis and the matching procedure.

### Study participants

We conducted a retrospective cohort study using Japanese longitudinal health checkup data. The participant selection process, including exclusion criteria, is illustrated in Fig 1. Participants comprised individuals who underwent the Specific Health Checkup at the Hokuriku Health Service Association in Toyama Prefecture, Japan, between April 2012 and March 2021. A total of 18,373 individuals aged 49–64 years (mean age: 53.0 ± 3.1 years) were initially identified.

**Fig 1.**
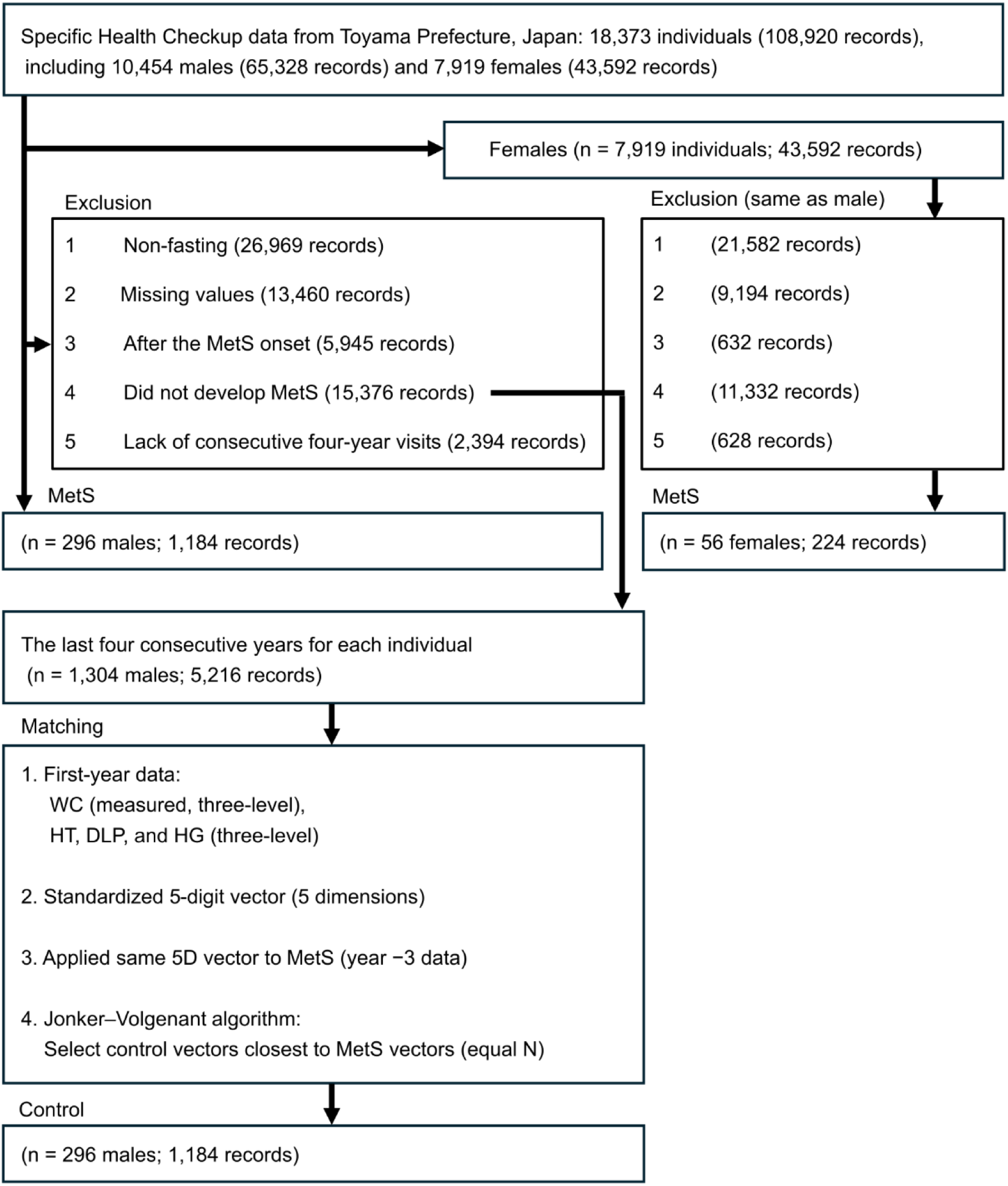
Flowchart of participant selection and matching procedure. Abbreviations: MetS, metabolic syndrome; WC, waist circumference; HT, hypertension; DLP, dyslipidemia; HG, hyperglycemia.

Due to sex-specific differences in the pathophysiology of MetS, analyses primarily focused on male participants, including comparisons with controls. Female data were included in the clustering analysis and analyzed separately from male participants.

Individuals who underwent non-fasting blood sampling or had missing values in any of the seven key variables—WC, SBP, DBP, TG, HDL-C, PG, or HbA1c—were also excluded. Additionally, records obtained after the onset of MetS, records from individuals who did not develop MetS during the study period, and records with missing values in any year from three years prior to onset to the year of onset were excluded. Ultimately, we selected 296 male participants diagnosed with MetS for inclusion in this study. As for females, 56 individuals were selected for the MetS group, and their results are presented in the Discussion section.

To identify suitable male controls, we first selected 15,376 records from non-MetS individuals and extracted the most recent four-year period from each individual, resulting in 1,304 four-year data sets. Second, for both MetS cases and controls, five variables were selected from the initial year of the four-year dataset per individual.

These included measured WC and three-level classification values of WC, HT, DLP, and HG, with the classification criteria described in S4 Table. WC was included as both a measured value and a three-level classification because OB is the central feature of MetS and provides complementary continuous and categorical information that improves the precision of matching. Each variable was standardized, and the five elements were treated as a five-dimensional vector for use in subsequent matching. Finally, matching was performed by calculating the Euclidean distance between the five-dimensional vectors of MetS cases and control. The Jonker–Volgenant algorithm [21] was applied to identify the closest matches. For each MetS case, an equal number of control individuals with the shortest Euclidean distances between the five-dimensional vectors were selected. Overlapping control candidates were not permitted. Ultimately, we selected 296 male participants without MetS as controls.

### Clustering analysis

For each MetS individual, the three-year pattern of four MetS components (WC, HT, DLP, and HG) was encoded as a 12-digit ternary code, as described in the “Encoding of MetS components” subsection. This encoded 12-dimensional vector was used for clustering analysis. To capture pre-onset longitudinal patterns, we used data from the three years preceding the onset year. The encoding procedure and structure of the input variables are illustrated in S1 Fig.

Hierarchical clustering was conducted using the Euclidean distance and Ward’s method [22]. To determine the optimal number of clusters, silhouette scores were calculated for cluster counts ranging from 2 to 10. Additionally, K-means clustering—a widely used alternative—was applied to the same dataset. Clustering performance was evaluated using the adjusted Rand index (ARI) [23].

### Group comparisons

Baseline characteristics, longitudinal features, and clinical outcomes were compared between MetS and control groups within each cluster. Comparisons were conducted using three comparison metrics: year −3, the change from year −3 to year 0, and year 0. In supplementary analyses, lifestyle factors were compared between MetS and control groups within each cluster, using two comparison metrics (year −3 to −1 and year −1 to 0). For variables measured between year −3 and −1, annual changes were estimated for each individual using simple linear regression, with time (in years) as the independent variable and each measurement as the dependent variable. This approach was used to improve the stability of individual trend estimates.

For continuous variables, normality was assessed using the Shapiro–Wilk test. When both groups met the assumption of normality, a two-sided unpaired Student’s *t*-test was used; otherwise, the Wilcoxon rank-sum test was applied. Although each MetS individual was matched with a control non-MetS individual, we considered that unpaired hypothesis tests were suitable because they were different persons. For comparisons of TyG-related indices across the clusters of the MetS group, the Kruskal–Wallis test was used at year 0, corresponding to the year of MetS onset. When significant differences were detected, Dunn’s test with Bonferroni correction was performed for post hoc multiple comparisons. For categorical variables, Fisher’s exact test was used when the minimum value of the expected cell counts were below 5; otherwise, the chi-square test was applied.

Continuous variables were presented as mean ± standard deviation or median with interquartile range, and categorical variables were reported as counts and percentages. A p-value < 0.05 was considered statistically significant.

Records with missing lifestyle information were handled as follows. Lifestyle analyses (S11–S14 Tables) were conducted using variable-specific denominators, excluding participants with missing values for each variable. Multiple linear regression analyses involving lifestyle variables excluded participants with any missing lifestyle data, as described in the next subsection.

### Multiple linear regression analysis

To evaluate factors associated with changes in WC, multiple linear regression analyses were conducted. Independent variables included age, self-reported smoking and alcohol consumption, engagement in lifestyle improvement efforts, and willingness to receive specific health guidance. These variables were selected as potential confounders of WC change. Annual changes were examined separately for two time periods (year −3 to −1 and year −1 to 0) to distinguish long-term trends from more recent changes. Analyses were conducted in both the combined MetS and control groups and the MetS group alone to evaluate overall associations as well as patterns characteristic to the MetS group.

Participants with any missing lifestyle information were excluded, and the resulting participants were as follows. For the period from year −3 to −1, data from 280 individuals in the MetS group (Cluster 1: 93; Cluster 2: 35; Cluster 3: 44; Cluster 4: 42; Cluster 5: 43; Cluster 6: 23) and 281 individuals in the control group (Cluster 1: 94; Cluster 2: 35; Cluster 3: 45; Cluster 4: 40; Cluster 5: 42; Cluster 6: 25) were analyzed. For the period from year −1 to 0, data from 279 individuals in the MetS group (Cluster 1: 90; Cluster 2: 36; Cluster 3: 44; Cluster 4: 42; Cluster 5: 44; Cluster 6: 23) and 282 individuals in the control group (Cluster 1: 94; Cluster 2: 35; Cluster 3: 46; Cluster 4: 40; Cluster 5: 42; Cluster 6: 25) were included.

Raw values were used for continuous variables. Reference categories for categorical variables are listed in S2 and S3 Tables. For cluster comparisons, Cluster 1 was used as the reference category. These coding choices were adopted to ensure consistent handling of variables across analyses.

### Calculation of TyG-related indices

TyG-WC and TyG-BMI were calculated as the product of TyG and WC, and TyG and BMI, respectively. TyG was defined as ln [TG (mg/dL) × PG (mg/dL) / 2], and BMI was calculated as body mass (kg) divided by height squared (m²).

### Software

Clustering, label matching, ARI evaluation, and estimation of annual change (year −3 to −1) were performed in Python (version 3.12.12) using the scikit-learn and scipy libraries. Statistical analyses, including hypothesis testing and multiple linear regression, were conducted in R (version 4.5.2) using base functions and the stats package.

### Ethics Statement

All procedures conducted in this study complied with the ethical standards of the 1964 Declaration of Helsinki and its subsequent amendments, as well as the Ethical Guidelines for Medical and Biological Research Involving Human Subjects issued by the Ministry of Health, Labour and Welfare of Japan. The study protocol was approved by the Ethics Committee of Toyama University Hospital (approval number: R2021070; approval date: 19 August 2021). All participants provided written informed consent for the use of their data in scientific research.

## Results

### Cohort characteristics and comparison of the six clusters

A total of 296 male participants with a median age of 55.0 years (interquartile range: 53.0–56.0) were included, yielding 1,184 records spanning four consecutive years. Through the matching procedure, 296 control individuals were selected (median age: 56.0 years; interquartile range: 54.0–58.0). The background characteristics are summarized in S5 Table.

The optimal number of clusters for hierarchical clustering was evaluated using the silhouette score (S2 Fig). Local maxima were observed at four and six clusters. Given the multifactorial nature of MetS, the six-cluster solution was considered appropriate to capture the heterogeneity of the study population. The dendrogram was cut at the corresponding height, resulting in the classification of participants into six clusters.

We also tested K-means clustering, but could not obtain consistent results. K-means clustering yielded the highest silhouette score with three clusters, followed by seven clusters, whereas the six-cluster solution showed a lower silhouette score. The ARI between the six-cluster solutions derived from hierarchical clustering and K-means clustering was 0.597, which is not high. In addition, the ARI between the six-cluster hierarchical solution and the seven-cluster K-means solution was 0.522. These results indicate that, in our dataset, the clustering outcomes derived from hierarchical clustering using Ward’s method were not fully consistent with those obtained using the K-means method. In this paper, we focused on the results of hierarchical clustering due to its deterministic nature.

The clinical characteristics of each cluster and the control group are summarized in Fig 2. The resulting clusters, defined by temporal patterns in MetS component changes, were as follows: Cluster 1 (n = 97, 32.8%), Cluster 2 (n = 36, 12.2%), Cluster 3 (n = 48, 16.2%), Cluster 4 (n = 44, 14.9%), Cluster 5 (n = 46, 15.5%), and Cluster 6 (n = 25, 8.4%). In the MetS group, the most frequent combinations of MetS components tended to shift between the early (year −3 to −1) and late (year −1 to 0) patterns, as shown in Fig 2. Cluster-specific patterns emerged—defined based on whether more than 50% of individuals in each cluster met the criteria for each MetS component: Cluster 1 showed early onset of OB and HT, followed by DLP; Cluster 2 began with OB and DLP, with later emergence of HT; Cluster 3 was marked by early OB and subsequent development of HT and HG; Cluster 4 exhibited early HT and DLP, with OB appearing later; Cluster 5 showed early HT, followed by OB, DLP, and HG; and Cluster 6 demonstrated early HT, DLP, and HG, with OB emerging later. At year 0, the prevalence of HT, DLP, and HG varied among clusters, with distinct combinations of these components observed in each cluster: in Clusters 1, 2, and 4, more than 50% individuals exhibited OB, HT, and DLP; Cluster 3 showed OB, HT, and HG; and Clusters 5 and 6 were characterized by the presence of all components—OB, HT, DLP, and HG. In contrast, the control group did not exhibit substantial temporal shifts in the most frequent patterns of MetS components, with the combinations observed at year 0 remaining largely similar to those seen at year −3.

**Fig 2.**
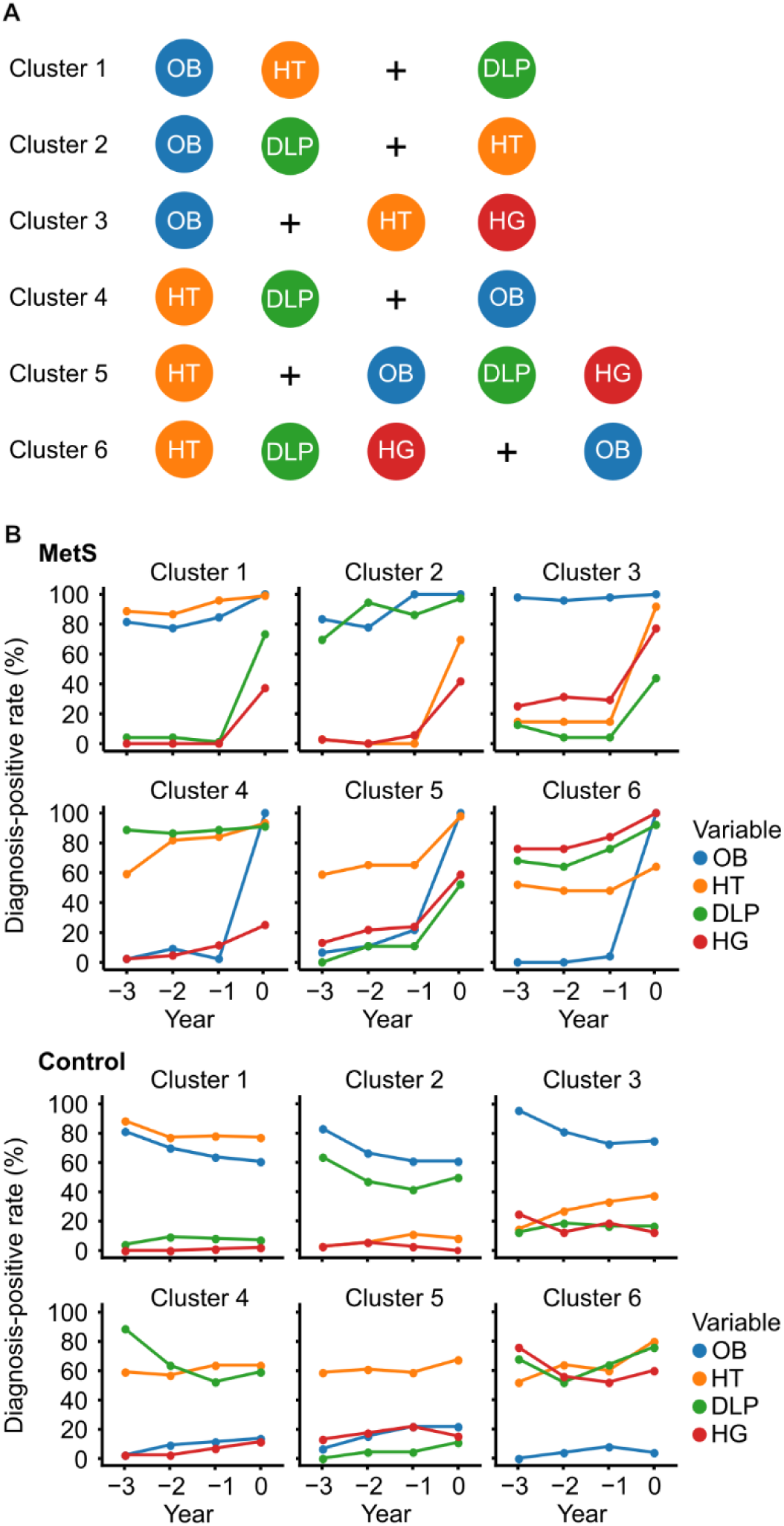
Summary of the characteristics of each cluster. (A) MetS component patterns observed in the early (year −3 to −1) and late (year −1 to 0) periods in the MetS group. For each cluster, MetS components are displayed side by side, with the early components shown on the left, followed by a ‘+’ symbol, and the late components on the right. The presence of each MetS component was determined based on whether more than 50% of individuals in the cluster met the criteria for that component, as assessed from the proportions shown in Fig 2B. (B) Diagnosis-positive rates for each MetS component—OB, HT, DLP, and HG—are plotted over time for clusters consisting of individuals diagnosed with MetS. Each cluster includes four color-coded line plots corresponding to the respective diagnostic components. In the upper panels, plots for the MetS group are arranged with clusters 1–3 in the top row and clusters 4–6 in the bottom row. In the lower panels, plots for the Control group are similarly arranged, with clusters 1–3 in the top row and clusters 4–6 in the bottom row. Abbreviations: MetS, metabolic syndrome; OB, abdominal obesity; HT, hypertension; DLP, dyslipidemia; HG, hyperglycemia.

### Comparison of clinical parameters across clusters and between MetS and control groups from year −3 to 0

To further explore the temporal dynamics underlying these component patterns, Figs 3 and 4 present the progression of related clinical variables from year −3 to 0 within each cluster. For variables associated with MetS components, a visual trend toward worsening was generally observed across clusters in the MetS group (Fig 3). Based on visual inspection of the annotated heatmap, certain variables in some clusters appeared to show a steeper increase between approximately year −1 and 0 compared to earlier years. Examples include TG in Cluster 1, SBP and DBP in Cluster 3, and WC in Clusters 4–6, which appeared to show more marked changes between year −1 and 0. In contrast to the MetS group, variables in the control group remained relatively stable, with no pronounced worsening observed across clusters throughout the observation period (Fig 4).

**Fig 3.**
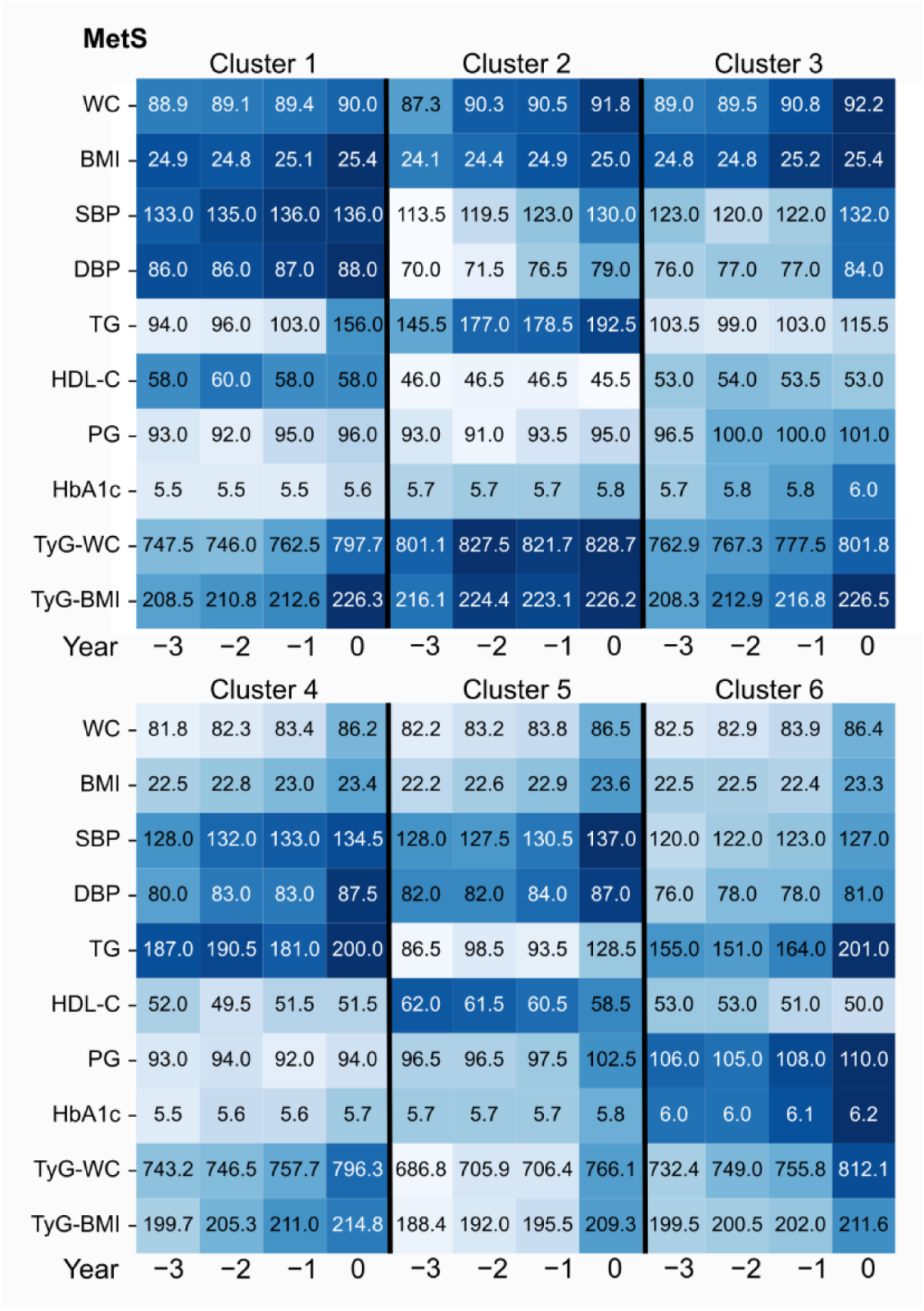
Heatmaps of clinical parameters from year −3 to 0 for Clusters 1–6 in the MetS group. Clusters 1–3 (top) and Clusters 4–6 (bottom) are displayed vertically. Each row represents a clinical variable, and each column corresponds to a specific year within each cluster. Continuous variables are shown as median values. Cluster names are shown above each cluster panel in the upper and lower heatmaps, and year labels are shown at the bottom. Color intensity reflects relative values standardized in a row-wise manner across all clusters in both the MetS and control groups, with darker colors indicating higher values and lighter colors indicating lower values. Abbreviations: MetS, metabolic syndrome; WC, waist circumference; BMI, body mass index; SBP, systolic blood pressure; DBP, diastolic blood pressure; TG, triglycerides; HDL-C, high-density lipoprotein cholesterol; PG, plasma glucose; HbA1c, glycated hemoglobin; TyG-WC, triglyceride-glucose index × waist circumference; TyG-BMI, triglyceride-glucose index × body mass index.

**Fig 4.**
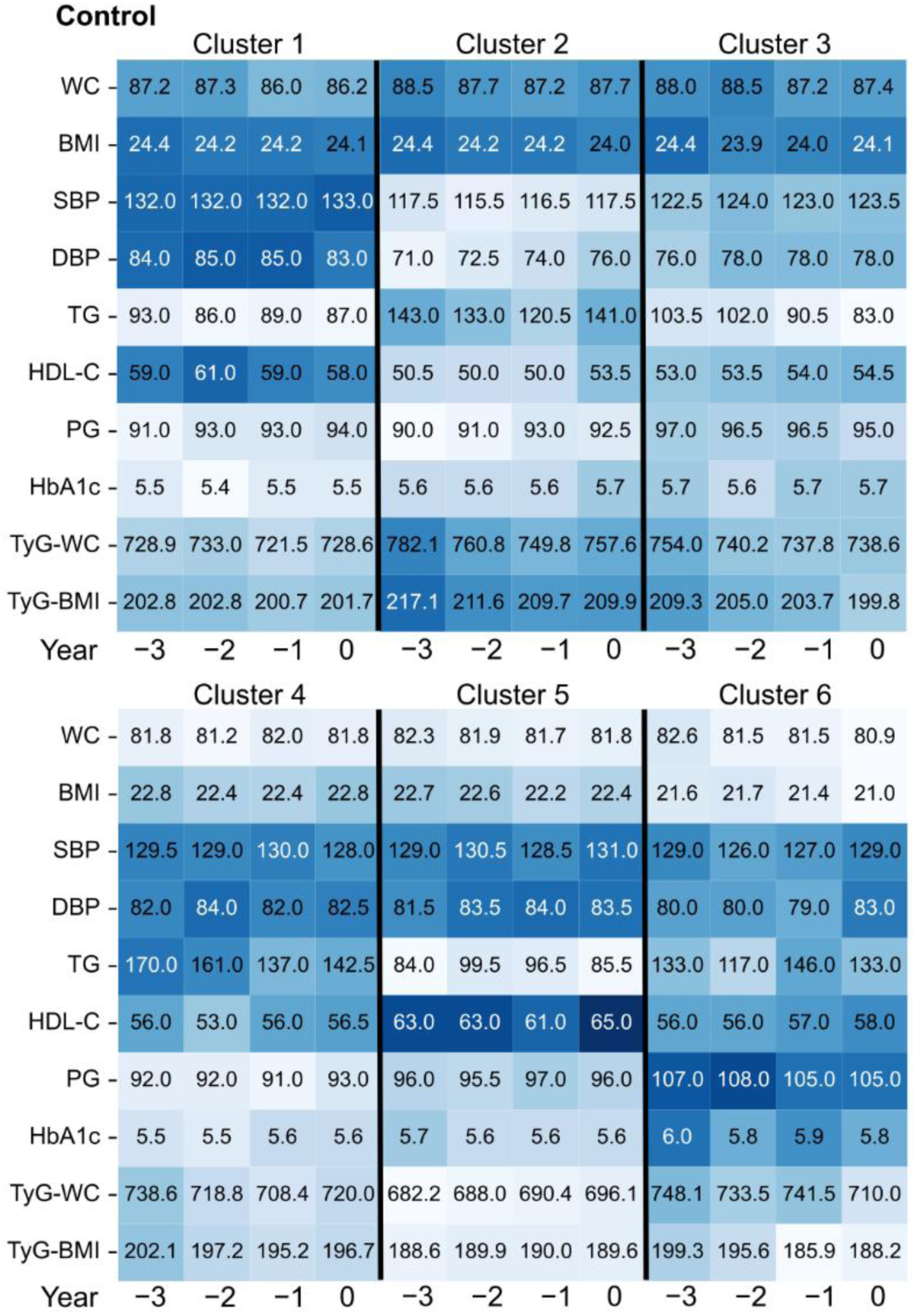
Heatmaps of clinical parameters from year −3 to 0 for Clusters 1–6 in the control group. Clusters 1–3 (top) and Clusters 4–6 (bottom) are displayed vertically. Each row represents a clinical variable, and each column corresponds to a specific year within each cluster. Continuous variables are shown as median values. Cluster names are shown above each cluster panel in the upper and lower heatmaps, and year labels are shown at the bottom. Color intensity reflects relative values standardized in a row-wise manner across all clusters in both the MetS and control groups, with darker colors indicating higher values and lighter colors indicating lower values. Abbreviations: WC, waist circumference; BMI, body mass index; SBP, systolic blood pressure; DBP, diastolic blood pressure; TG, triglycerides; HDL-C, high-density lipoprotein cholesterol; PG, plasma glucose; HbA1c, glycated hemoglobin; TyG-WC, triglyceride-glucose index × waist circumference; TyG-BMI, triglyceride-glucose index × body mass index.

To complement these findings, S3 and S4 Figs illustrate how combinations of MetS components were distributed over time across clusters, providing a categorical perspective on component patterns in both the MetS and control groups. In the MetS group, distinct temporal patterns of component combinations were observed among clusters (S3 Fig). From year −3 to −1, Cluster 1 was characterized by the dominant combination of OB and HT, which shifted to the combination of OB, HT, and DLP at year 0. Cluster 2 showed the most frequent combination of OB and DLP from year −3 to −1, shifting to the combination of OB, HT, and DLP at year 0. In Cluster 3, OB alone was the most common pattern from year −3 to −1, shifting to the combination of OB, HT, and HG at year 0. Cluster 4 was characterized by the combination of HT and DLP from year −3 to −1, with a shift to the combination of OB, HT, and DLP at year 0. In Cluster 5, HT alone was the most prevalent pattern from year −3 to −1, shifting to the combination of OB, HT, and HG at year 0, followed by the combination of OB, HT, and DLP. In Cluster 6, the combination of HT, DLP, and HG was the commonly observed combination from year −3 to −1, which shifted to the combination of OB, HT, DLP, and HG at year 0. In contrast, the control group exhibited relatively stable patterns from year −3 to 0, often maintaining the same combinations observed in the corresponding MetS clusters from year −3 to −1 (S4 Fig). In summary, the pattern of MetS component combination was relatively stable until year −1 in all clusters, and the MetS group experienced remarkable changes in the last year before the onset of MetS.

Clinical characteristics of the MetS and control groups at year −3 for each cluster are presented in S6 and S7 Tables. No significant differences were observed between the MetS and control groups within most clusters except for age. Modest group differences were observed in Clusters 1, 2, 3, and 6, where the MetS group tended to be younger. In Cluster 2, HDL-C levels were lower in the MetS group compared to controls.

Changes in clinical parameters observed between year −3 and −1, or between year −1 and 0, are presented in Tables 1 and 2. Table 1 summarizes data for Clusters 1–3, while Table 2 presents the corresponding data for Clusters 4–6. Detailed definitions and descriptions of selected variables are provided in S8 Table. As shown in Tables 1 and 2, annual WC change was significantly greater in all clusters of the MetS group between year −3 and −1, and remained significant between year −1 and 0 in Clusters 1, 4, 5, and 6. BMI exhibited a broadly similar pattern across both time periods.

**Table 1.**
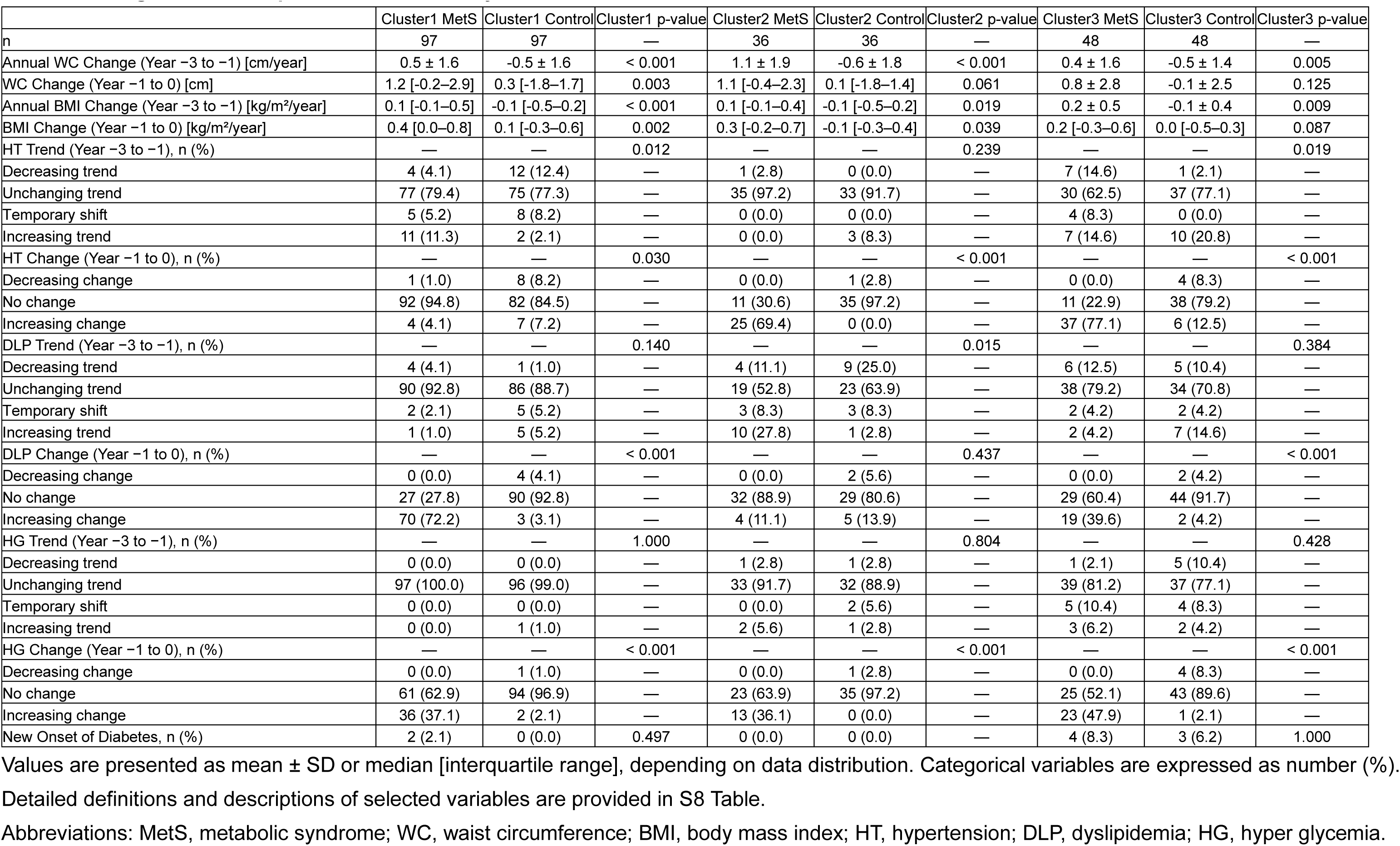
Changes in clinical parameters between year −3 and 0 in Clusters 1–3.

|  | Cluster1 MetS | Cluster1 Control | Cluster1 p-value | Cluster2 MetS | Cluster2 Control | Cluster2 p-value | Cluster3 MetS | Cluster3 Control | Cluster3 p-value |
| --- | --- | --- | --- | --- | --- | --- | --- | --- | --- |
| n | 97 | 97 | — | 36 | 36 | — | 48 | 48 | — |
| Annual WC Change (Year -3 to -1) [cm/year] | 0.5 ± 1.6 | -0.5 ± 1.6 | < 0.001 | 1.1 ± 1.9 | -0.6 ± 1.8 | < 0.001 | 0.4 ± 1.6 | -0.5 ± 1.4 | 0.005 |
| WC Change (Year -1 to 0) [cm] | 1.2 [-0.2–2.9] | 0.3 [-1.8–1.7] | 0.003 | 1.1 [-0.4–2.3] | 0.1 [-1.8–1.4] | 0.061 | 0.8 ± 2.8 | -0.1 ± 2.5 | 0.125 |
| Annual BMI Change (Year -3 to -1) [kg/m<sup>2</sup>/year] | 0.1 [-0.1–0.5] | -0.1 [-0.5–0.2] | < 0.001 | 0.1 [-0.1–0.4] | -0.1 [-0.5–0.2] | 0.019 | 0.2 ± 0.5 | -0.1 ± 0.4 | 0.009 |
| BMI Change (Year -1 to 0) [kg/m<sup>2</sup>/year] | 0.4 [0.0–0.8] | 0.1 [-0.3–0.6] | 0.002 | 0.3 [-0.2–0.7] | -0.1 [-0.3–0.4] | 0.039 | 0.2 [-0.3–0.6] | 0.0 [-0.5–0.3] | 0.087 |
| HT Trend (Year -3 to -1), n (%) | — | — | 0.012 | — | — | 0.239 | — | — | 0.019 |
| Decreasing trend | 4 (4.1) | 12 (12.4) | — | 1 (2.8) | 0 (0.0) | — | 7 (14.6) | 1 (2.1) | — |
| Unchanging trend | 77 (79.4) | 75 (77.3) | — | 35 (97.2) | 33 (91.7) | — | 30 (62.5) | 37 (77.1) | — |
| Temporary shift | 5 (5.2) | 8 (8.2) | — | 0 (0.0) | 0 (0.0) | — | 4 (8.3) | 0 (0.0) | — |
| Increasing trend | 11 (11.3) | 2 (2.1) | — | 0 (0.0) | 3 (8.3) | — | 7 (14.6) | 10 (20.8) | — |
| HT Change (Year -1 to 0), n (%) | — | — | 0.030 | — | — | < 0.001 | — | — | < 0.001 |
| Decreasing change | 1 (1.0) | 8 (8.2) | — | 0 (0.0) | 1 (2.8) | — | 0 (0.0) | 4 (8.3) | — |
| No change | 92 (94.8) | 82 (84.5) | — | 11 (30.6) | 35 (97.2) | — | 11 (22.9) | 38 (79.2) | — |
| Increasing change | 4 (4.1) | 7 (7.2) | — | 25 (69.4) | 0 (0.0) | — | 37 (77.1) | 6 (12.5) | — |
| DLP Trend (Year -3 to -1), n (%) | — | — | 0.140 | — | — | 0.015 | — | — | 0.384 |
| Decreasing trend | 4 (4.1) | 1 (1.0) | — | 4 (11.1) | 9 (25.0) | — | 6 (12.5) | 5 (10.4) | — |
| Unchanging trend | 90 (92.8) | 86 (88.7) | — | 19 (52.8) | 23 (63.9) | — | 38 (79.2) | 34 (70.8) | — |
| Temporary shift | 2 (2.1) | 5 (5.2) | — | 3 (8.3) | 3 (8.3) | — | 2 (4.2) | 2 (4.2) | — |
| Increasing trend | 1 (1.0) | 5 (5.2) | — | 10 (27.8) | 1 (2.8) | — | 2 (4.2) | 7 (14.6) | — |
| DLP Change (Year -1 to 0), n (%) | — | — | < 0.001 | — | — | 0.437 | — | — | < 0.001 |
| Decreasing change | 0 (0.0) | 4 (4.1) | — | 0 (0.0) | 2 (5.6) | — | 0 (0.0) | 2 (4.2) | — |
| No change | 27 (27.8) | 90 (92.8) | — | 32 (88.9) | 29 (80.6) | — | 29 (60.4) | 44 (91.7) | — |
| Increasing change | 70 (72.2) | 3 (3.1) | — | 4 (11.1) | 5 (13.9) | — | 19 (39.6) | 2 (4.2) | — |
| HG Trend (Year -3 to -1), n (%) | — | — | 1.000 | — | — | 0.804 | — | — | 0.428 |
| Decreasing trend | 0 (0.0) | 0 (0.0) | — | 1 (2.8) | 1 (2.8) | — | 1 (2.1) | 5 (10.4) | — |
| Unchanging trend | 97 (100.0) | 96 (99.0) | — | 33 (91.7) | 32 (88.9) | — | 39 (81.2) | 37 (77.1) | — |
| Temporary shift | 0 (0.0) | 0 (0.0) | — | 0 (0.0) | 2 (5.6) | — | 5 (10.4) | 4 (8.3) | — |
| Increasing trend | 0 (0.0) | 1 (1.0) | — | 2 (5.6) | 1 (2.8) | — | 3 (6.2) | 2 (4.2) | — |
| HG Change (Year -1 to 0), n (%) | — | — | < 0.001 | — | — | < 0.001 | — | — | < 0.001 |
| Decreasing change | 0 (0.0) | 1 (1.0) | — | 0 (0.0) | 1 (2.8) | — | 0 (0.0) | 4 (8.3) | — |
| No change | 61 (62.9) | 94 (96.9) | — | 23 (63.9) | 35 (97.2) | — | 25 (52.1) | 43 (89.6) | — |
| Increasing change | 36 (37.1) | 2 (2.1) | — | 13 (36.1) | 0 (0.0) | — | 23 (47.9) | 1 (2.1) | — |
| New Onset of Diabetes, n (%) | 2 (2.1) | 0 (0.0) | 0.497 | 0 (0.0) | 0 (0.0) | — | 4 (8.3) | 3 (6.2) | 1.000 |

Values are presented as mean ± SD or median [interquartile range], depending on data distribution. Categorical variables are expressed as number (%).
Detailed definitions and descriptions of selected variables are provided in S8 Table.
Abbreviations: MetS, metabolic syndrome; WC, waist circumference; BMI, body mass index; HT, hypertension; DLP, dyslipidemia; HG, hyper glycemia.

**Table 2.**
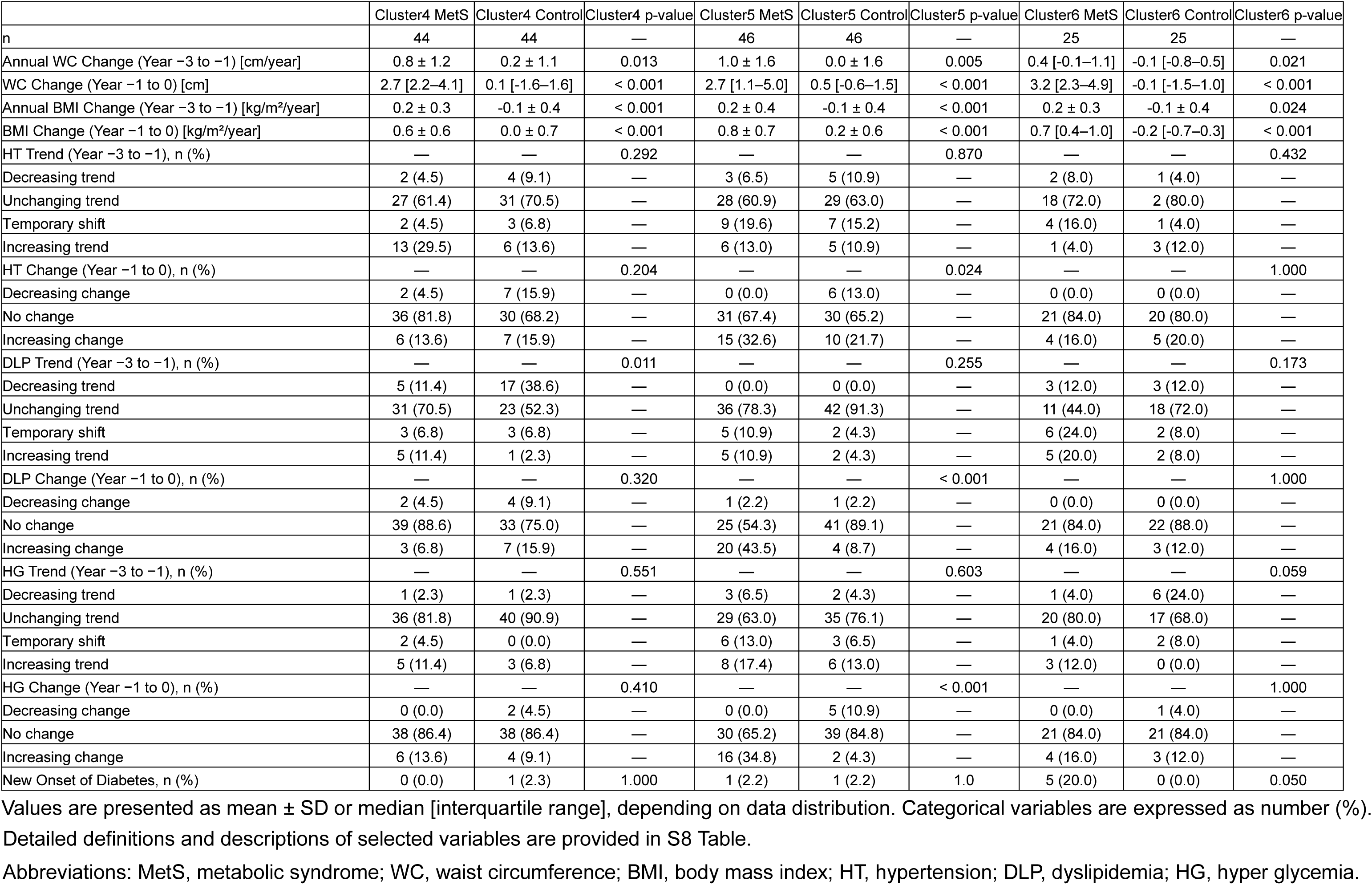
Changes in clinical parameters between year −3 and 0 in Clusters 4–6.

Regarding MetS components, between year −3 and −1, Cluster 1 in the MetS group showed a significantly different distribution of HT change patterns compared to the control group, with a tendency toward increase, while the control group exhibited a trend toward improvement. Clusters 2 and 4 in the MetS group tended to show increases in DLP during this period, while the control group showed a trend towards improvement. In Cluster 3, HT showed a slower progression in the MetS group compared to the control group. Cluster 4 was described above. Cluster 5 did not show clear differences compared to the control group. In Cluster 6, the MetS group tended to show an increase in HG, while the control group showed slight improvement in this component. Between year −1 and 0, Clusters 1, 2, 3, and 5 in the MetS group demonstrated significant increases in at least two components among HT, DLP, and HG. Clusters 4 and 6 exhibited no significant changes during this latter period.

Between year −3 and 0, the MetS group in Cluster 6 showed a trend toward a higher incidence of new-onset diabetes compared to the control group.

S9 and S10 Tables present the clinical characteristics of the MetS and control groups at year 0 for each cluster. S9 Table summarizes data for Clusters 1–3, while S10 Table presents the corresponding data for Clusters 4–6. Across all clusters, the MetS group exhibited significantly higher values in WC compared to the control group. A similar trend was observed for BMI, although the difference was not statistically significant in Cluster 2. In the MetS group, the proportion of individuals with a BMI ≥ 25 was 52.8%–59.8% in Clusters 1–3 and 4.5%–24.0% in Clusters 4–6, with all individuals having a WC ≥ 85 cm, by definition. The MetS group tended to have a greater number of MetS components, with statistically significant differences in the distribution observed in Clusters 1–5, and a similar trend was observed in Cluster 6. TyG-WC and TyG-BMI indices were consistently elevated in the MetS group across all clusters.

S11–S14 Tables present lifestyle-related characteristics of the MetS and control groups. Definitions and descriptions of the lifestyle variables are provided in S1–S3 Tables. Except for a significant difference observed in alcohol-related variables in Cluster 4, no statistically significant differences were found between the MetS and control groups in any of the lifestyle-related parameters analyzed during either period.

### Temporal predictors of WC change and TyG-related indices across clusters at year 0

Cluster-specific changes in WC are illustrated in S5 Fig. To explore temporal differences in predictors of WC change, multiple linear regression analyses were conducted separately for the periods from year −3 to −1 and from year −1 to 0. S6 and S7 Figs present the results of these analyses. In both periods, smoking cessation and belonging to the MetS group were significantly associated with annual changes in WC. During the period from year −1 to 0, Clusters 4, 5, and 6 were significantly associated with greater increases in WC compared to Cluster 1.

In addition to the temporal analysis of predictors of WC change, TyG-WC and TyG-BMI were compared across clusters at year 0. Fig 5 illustrates the distribution of TyG-WC and TyG-BMI across clusters at year 0. At this time point (the year of MetS onset), significant differences in both indices were observed among MetS clusters, as determined by the Kruskal–Wallis rank sum test (TyG-WC: *χ²* = 42.914, df = 5, p < 0.001; TyG-BMI: *χ²* = 36.582, df = 5, p < 0.001). Post hoc multiple comparisons using Dunn’s test revealed that, for TyG-WC, Cluster 2 showed significantly higher values compared to Clusters 4 and 5. Additionally, Clusters 1, 3, and 6 had significantly higher values than Cluster 5. Regarding TyG-BMI, Clusters 1 and 2 exhibited significantly higher values than Clusters 4 and 5, and Cluster 3 also showed significantly higher values than Cluster 5.

**Fig 5.**
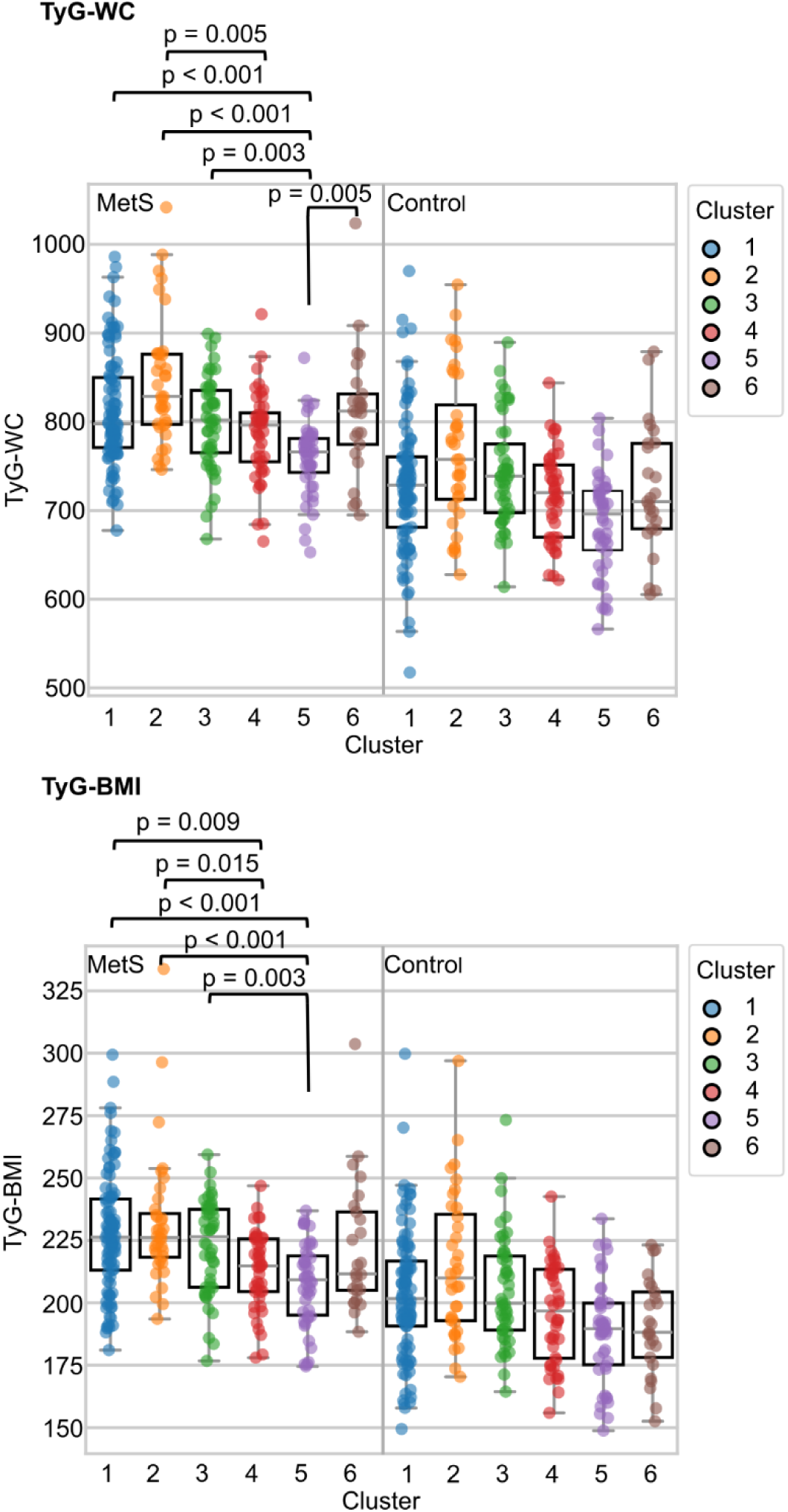
Distribution of TyG-WC and TyG-BMI across clusters at year 0. P-values displayed above the box plots represent Bonferroni-adjusted values for multiple comparisons among clusters within the MetS group. Abbreviations: MetS, metabolic syndrome; WC, waist circumference; BMI, body mass index; TyG-WC, triglyceride-glucose index × waist circumference; TyG-BMI, triglyceride-glucose index × body mass index.

## Discussion

This study applied hierarchical clustering to Japanese health checkup data from 296 male individuals who developed MetS, capturing the trajectories of MetS components during the three years preceding diagnosis. Based on these trajectories, participants were classified into six clusters. As summarized in Fig 2, the six clusters showed differences in the frequent patterns between the early (year −3 to −1) and late (year −1 to 0) periods. Cluster 1 (n = 97, 32.8%) showed early OB and HT, followed by DLP. Cluster 2 (n = 36, 12.2%) showed early OB and DLP, followed by HT. Cluster 3 (n = 48, 16.2%) showed early OB, followed by HT and HG. Cluster 4 (n = 44, 14.9%) showed early HT and DLP, with later OB. Cluster 5 (n = 46, 15.5%) showed early HT, followed by OB, DLP, and HG. Cluster 6 (n = 25, 8.4%) showed early HT, DLP, and HG, followed by OB. In contrast, no clear changes were observed in controls. These clusters revealed multiple patterns of progression, suggesting that the clustering approach provided a useful framework for characterizing heterogeneous pathways to MetS, which are often difficult to describe because of their inherent complexity.

The clinical relevance of the identified clusters may be supported by prior evidence, such as a longitudinal study that identified a combination of OB, HT, and HG as strong predictors of cardiovascular disease and mortality [24]. Although this study did not evaluate the long-term outcomes of each cluster, TyG-WC and TyG-BMI—recognized as promising biomarkers for screening and predicting cardiovascular disease and insulin resistance [25–28]—showed significant differences across MetS clusters. This suggests that each cluster may be associated with varying levels of cardiometabolic risk, with Cluster 2 being the highest and Custer 5 the lowest. In addition to TyG-related indices, Clusters 4–6 include individuals who exhibit cardiometabolic abnormalities that may precede visceral fat accumulation—a pattern that could suggest an underlying genetic predisposition and a longer duration of metabolic dysfunction. Notably, a large meta-analysis has shown that metabolically unhealthy individuals with normal weight may have a higher risk of all-cause mortality and/or cardiovascular events than their overweight or obese counterparts [29]. Therefore, individuals in Clusters 4–6 may also have a large risk of cardiovascular events. In addition, although all MetS clusters showed significantly greater WC changes compared to the control group, Clusters 4–6 specifically included individuals with rapid WC increases. These findings suggest that even non-obese individuals should pay attention to clinical outcomes, along with sudden increases in WC that may trigger the onset of MetS.

Although the primary analysis of this study focused on male participants, we additionally performed hierarchical clustering on female participants using the same method to evaluate pre-onset patterns of MetS components (S8 Fig). Three clusters were determined based on silhouette scores and clinical interpretability. In contrast to males, HG appeared more frequently in females. This tendency was observed in the comparisons between female Cluster 1 and male Cluster 2, between female Cluster 2 and male Cluster 1, and between female Cluster 3 and male Cluster 5 at one or more corresponding time points within the observation period. In addition, when comparing female Cluster 3 with male Cluster 5, OB appeared more frequently in females between year −3 and −1. Additionally, the progression of MetS components appeared more gradual in females over the period from year −3 to 0, especially in the comparison between female Cluster 1 and male Cluster 2. A previous study reported that age-related increases in MetS components have been observed in both sexes, with HT and HG showing pronounced rise, especially in females [30]. In addition, OB has been characterized as a more common feature of MetS in females than in males [30]. This sex difference may be associated with postmenopausal hormonal dysregulation, which predisposes females to visceral fat accumulation [31]. Another study suggests that, although the speed of MetS development is expected to be higher in females than in males—reflecting the elevated MetS prevalence in postmenopausal females—model-based predictions indicate that females are more likely to show improvement in MetS components, and may show a slower progression of MetS [16]. Although our detailed analysis focused on males, previous studies have reported a higher prevalence of MetS among females in postmenopausal age groups [16,30,31], indicating the need to focus on sex-specific investigations.

Our findings revealed a younger age among individuals who developed MetS, diverging from previous reports of age-related increases in MetS prevalence [32,33]. This discrepancy may be partly explained by the analysis method. For each non-MetS individual, the most recent (or oldest) consecutive four-year period was extracted before matching. This procedure may have introduced an older age bias in the control group. In addition, a previous study conducted in Japanese cohorts has reported that, based on criteria requiring obesity, the prevalence of MetS in males generally increases with age but tends to plateau around 50–59 years [34]. Another study reported that, although the prevalence of HT and HG increases with age in males, OB and high TG levels do not show a similar age-related increase [30].

Therefore, the observed age distribution may partly reflect Japanese population trends, possibly influenced by obesity-based criteria, rather than solely methodological bias.

Among lifestyle factors, smoking cessation stood out as significantly associated with WC gain. Previous studies have shown that smoking cessation is associated with weight gain and unfavorable changes in metabolic profiles [35]. However, cardiovascular and mortality benefits of smoking cessation outweigh the harms associated with weight gain, especially beyond the initial few years [36]. To maximize these benefits, personalized weight management strategies should be considered to mitigate increases in WC following smoking cessation. In addition to smoking cessation and the lifestyle factors analyzed in this study, various other factors—including physical activity, diet, sleep, sedentary behavior, social contact, and working time—have been reported to be associated with MetS [37]. As suggested by these studies, we expected that lifestyle-related risks might differ between MetS and control groups within clusters, enabling cluster-specific interventions. However, no clinically meaningful differences were observed in lifestyle factors related to diet, physical activity or sleep between the MetS and control groups within each cluster. Therefore, these factors were collectively examined under the broader category of ‘lifestyle efforts’. Further investigations using larger datasets or alternative analytical approaches may help identify lifestyle-related risk patterns.

The pathogenesis of MetS involves several complex pathways and remains incompletely understood [38]. The interplay among factors such as insulin resistance, chronic inflammation, endothelial dysfunction, oxidative stress, the sympathetic nervous system, and the renin–angiotensin–aldosterone system exerts various adverse effects, including disturbances in lipid and glucose metabolism and harmful effects on the vascular system [3]. Beyond these molecular mechanisms, various factors—including ethnicity, sex, age, genetic predisposition, physical activity, and nutrition—also influence the pathophysiology of MetS [3,38]. The multifaceted nature and variability in clinical characteristics suggest the need to identify patterns that lead to MetS. In future research, the clustering approach presented in this study may help generate insights that contribute to a deeper understanding of the underlying pathophysiology associated with susceptibility profiles related to MetS components.

In addition, the findings derived from this approach offer important insights for the development of tailored prevention strategies for MetS. Across all clusters, individuals with increasing WC warrant particular attention. Specific considerations include: In Cluster 1, individuals with coexisting OB and HT may include those at increased risk of developing DLP, with persistent HT being one of the characteristics observed among those at risk; in Cluster 2, characterized by the coexistence of OB and DLP, unresolved DLP may represent a feature associated with the development of HT; in Cluster 3, continued OB is associated with increased risk of HT and HG; in Cluster 4, persistent DLP may indicate an elevated risk for progression to MetS; In Cluster 5, multiple MetS components emerge sequentially in close succession; and in Cluster 6, individuals at higher risk of diabetes onset may be included.

Additionally, Clusters 4, 5, and 6 require vigilance for rapid increases in WC. MetS components are closely interrelated and may independently or synergistically contribute to its progression [38]. These insights reinforce the importance of early intervention, as some factors show greater potential for improvement. For example, DLP responds more favorably to treatment compared to other MetS components [16], and that individuals without obesity tend to have a greater potential for improvement in MetS [39]. In contrast to these favorable indications, males aged ≥ 50 years with two MetS components (with obesity included in the count) have been reported to have a risk of progression of additional components within five years, in some cases exceeding 50% [15]. Furthermore, as suggested by this study, additional components may emerge rapidly within a short timeframe. Therefore, to prevent further progression, regular monitoring of other MetS components based on cluster-specific patterns is recommended. In particular, closer monitoring of WC and shorter follow-up intervals when increases occur may enhance the effectiveness of preventive strategies.

This study has several limitations that should be considered when interpreting the findings. First, differences between the hierarchical clustering results and those from K-means suggest inconsistency across methods. Given the varying assumptions and sensitivity of clustering algorithms to factors like shape, density, and distribution, such variation is expected. In this study, hierarchical clustering was selected due to its appropriateness for its deterministic nature, which avoids reliance on random initialization. However, for larger-scale datasets, K-means may offer greater computational efficiency and could be a more suitable alternative in future applications [4,40]. Second, the analysis was based on a Japanese male population, which may limit generalizability to other ethnic groups. In addition, Japanese have a lower average BMI and a lower prevalence of obesity compared with Caucasians, which also limits the generalizability of our findings. Third, the absence of socioeconomic and genetic information prevented us from examining differences in these factors across clusters, and the resulting bias is difficult to determine. These additional data may have helped refine the interpretation of the clusters. Fourth, the follow-up period was relatively short (4 years), limiting the ability to observe long-term outcomes such as cardiovascular disease or stroke across clusters. Although we also conducted analyses using a 6-year period (S9 Fig), changes in males remained relatively large from year −1 to 0. However, due to the reduced number of individuals and the duration still being insufficient for evaluating long-term outcomes, the extended period offered little additional benefit. Therefore, in this study, we focused on changes in males from year −1 to 0 and selected the 4-year period to maximize the number of individuals included in the analysis. Fifth, the restricted age range of participants may have influenced cluster composition, and clusters might differ if a broader age distribution were included. Overall, the clustering approach has potential for broader application, supported by similar patterns observed across different follow-up durations and by findings in female participants that were consistent with characteristics reported in previous studies. Nevertheless, the interpretation of the results remains influenced by the characteristics of the study population.

In conclusion, this study revealed multiple pre-onset pathways to MetS and demonstrated the utility of hierarchical clustering on longitudinal health checkup data. We identified six patterns of MetS development, characterized by different order of the emergence of MetS components. More frequent monitoring of MetS components could support the development of more personalized intervention strategies for each cluster. In addition, an increase in WC was observed in all clusters, suggesting that changes in WC are a common predictor of MetS—even among non-obese individuals. These findings indicate the importance of tracking WC changes over time, along with measuring WC levels. Taken together, given the inherent complexity of MetS and the difficulty of describing its developmental pathways, this approach provides a flexible framework for capturing the heterogeneity of pathways leading to MetS and may allow more targeted early intervention strategies tailored to population-specific patterns.

## Data Availability

The datasets generated during and/or analyzed in the current study are available from the corresponding author upon a substantiated request.

## Acknowledgements

We sincerely thank all the participants who underwent health checkups at the Hokuriku Health Service Association in Toyama Prefecture, Japan, for their cooperation in this study. We are also grateful to the entire staff of the Hokuriku Health Service Association for their dedicated support during data collection. We would like to thank all members of the Research Center for Pre-Disease Science, University of Toyama, Toyama, Japan, for their valuable input and encouragement throughout the research process.

## Funding

This research was supported by JST Moonshot R&D Grant Number JPMJMS2021. The funder had no role in the study design, data collection, data analysis, decision to publish, or preparation of the manuscript.

## Author contributions

S.S. was involved in the study design, data analysis, and interpretation. Y.N. and T.Y. were responsible for data provision and contributed to data acquisition. M.O. and K.T. provided supervision and guidance throughout the research process. All authors reviewed and approved the final manuscript.

## Competing interests

The authors declare no competing interests.

## Supporting information

**S1 Table. Binary coding of questionnaire responses.** This table provides binary coding definitions for self-administered questionnaire items used in lifestyle characteristics (S11–S14 Tables) and multivariable regression analyses.

**S2 Table. Variable definitions (year −3 to −1).** Variables derived from questionnaire responses (S1 Table), collected during the period from year −3 to −1.

**S3 Table. Variable definitions (year −1 to 0).** Variables derived from questionnaire responses (S1 Table), collected during the period from year −1 to 0.

**S4 Table. Criteria for three-level classification of MetS components.** Abbreviations: MetS, metabolic syndrome; WC, waist circumference; SBP, systolic blood pressure; DBP, diastolic blood pressure; TG, triglycerides; HDL-C, high-density lipoprotein cholesterol; FPG, fasting plasma glucose; HbA1c, glycated hemoglobin.

**S5 Table. Background characteristics of study participants (year −3).** All continuous variables were non-normally distributed and are presented as median [interquartile range]. Categorical variables are expressed as number (%). “MetS component, n (%)” indicates the number of components meeting the diagnostic criteria for MetS among the following: HT, DLP, HG, excluding OB. Abbreviations: MetS, metabolic syndrome; WC, waist circumference; BMI, body mass index; SBP, systolic blood pressure; DBP, diastolic blood pressure; HT, hypertension; TG, triglycerides; HDL-C, high-density lipoprotein cholesterol; DLP, dyslipidemia; PG, plasma glucose; HbA1c, glycated hemoglobin; HG, hyperglycemia; TyG-WC, triglyceride-glucose index × waist circumference; TyG-BMI, triglyceride-glucose index × body mass index.

**S6 Table. Clinical characteristics of MetS and control groups at year −3 (Clusters 1–3).** Values are presented as mean ± SD or median [interquartile range], depending on data distribution. Categorical variables are expressed as number (%). “MetS component, n (%)” indicates the number of components meeting the diagnostic criteria for MetS among the following: HT, DLP, HG, excluding OB. Abbreviations: MetS, metabolic syndrome; WC, waist circumference; BMI, body mass index; SBP, systolic blood pressure; DBP, diastolic blood pressure; HT, hypertension; TG, triglycerides; HDL-C, high-density lipoprotein cholesterol; DLP, dyslipidemia; PG, plasma glucose; HbA1c, glycated hemoglobin; HG, hyper glycemia; TyG-WC, triglyceride-glucose index × waist circumference; TyG-BMI, triglyceride-glucose index × body mass index.

**S7 Table. Clinical characteristics of MetS and control groups at year −3 (Clusters 4–6).** Values are presented as mean ± SD or median [interquartile range], depending on data distribution. Categorical variables are expressed as number (%). “MetS component, n (%)” indicates the number of components meeting the diagnostic criteria for MetS among the following: HT, DLP, HG, excluding OB. Abbreviations: MetS, metabolic syndrome; WC, waist circumference; BMI, body mass index; SBP, systolic blood pressure; DBP, diastolic blood pressure; HT, hypertension; TG, triglycerides; HDL-C, high-density lipoprotein cholesterol; DLP, dyslipidemia; PG, plasma glucose; HbA1c, glycated hemoglobin; HG, hyper glycemia; TyG-WC, triglyceride-glucose index × waist circumference; TyG-BMI, triglyceride-glucose index × body mass index.

**S8 Table. Detailed definitions and descriptions of selected variables.** “Yes” indicates that the individual met the diagnostic criterion for the corresponding MetS component (HT, DLP, or HG) in that year. “No” indicates that the criterion was not met. Abbreviations: MetS, metabolic syndrome; HT, hypertension; DLP, dyslipidemia; HG, hyper glycemia.

**S9 Table. Clinical characteristics of MetS and control groups at year 0 in Clusters 1–3.** Values are presented as mean ± SD or median [interquartile range], depending on data distribution. Categorical variables are expressed as number (%). “MetS component, n (%)” indicates the number of components meeting the diagnostic criteria for MetS among the following: HT, DLP, HG, excluding OB. Abbreviations: MetS, metabolic syndrome; WC, waist circumference; BMI, body mass index; SBP, systolic blood pressure; DBP, diastolic blood pressure; HT, hypertension; TG, triglycerides; HDL-C, high-density lipoprotein cholesterol; DLP, dyslipidemia; PG, plasma glucose; HbA1c, glycated hemoglobin; HG, hyperglycemia; TyG-WC, triglyceride-glucose index × waist circumference; TyG-BMI, triglyceride-glucose index × body mass index.

S10 Table. Clinical characteristics of MetS and control groups at year 0 in Clusters 4–6.

Values are presented as mean ± SD or median [interquartile range], depending on data distribution. Categorical variables are expressed as number (%). “MetS component, n (%)” indicates the number of components meeting the diagnostic criteria for MetS among the following: HT, DLP, HG, excluding OB. Abbreviations: MetS, metabolic syndrome; WC, waist circumference; BMI, body mass index; SBP, systolic blood pressure; DBP, diastolic blood pressure; HT, hypertension; TG, triglycerides; HDL-C, high-density lipoprotein cholesterol; DLP, dyslipidemia; PG, plasma glucose; HbA1c, glycated hemoglobin; HG, hyperglycemia; TyG-WC, triglyceride-glucose index × waist circumference; TyG-BMI, triglyceride-glucose index × body mass index.

**S11 Table. Lifestyle characteristics of MetS and control groups in Clusters 1–3 (year −3 to −1).** All continuous variables were non-normally distributed and are presented as median [interquartile range]. Categorical variables are expressed as number (%). For each variable, participants with missing values were excluded from the variable-specific denominators. The group-specific sample sizes shown in the row labeled “n” represent the total number of participants in each group, including those with missing lifestyle data.

**S12 Table. Lifestyle characteristics of MetS and control groups in Clusters 1–3 (year −1 to 0).** Categorical variables are expressed as number (%). For each variable, participants with missing values were excluded from the variable-specific denominators. The group-specific sample sizes shown in the row labeled “n” represent the total number of participants in each group, including those with missing lifestyle data.

**S13 Table. Lifestyle characteristics of MetS and control groups in Clusters 4–6 (year −3 to −1).** All continuous variables were non-normally distributed and are presented as median [interquartile range]. Categorical variables are expressed as number (%). For each variable, participants with missing values were excluded from the variable-specific denominators. The group-specific sample sizes shown in the row labeled “n” represent the total number of participants in each group, including those with missing lifestyle data.

**S14 Table. Lifestyle characteristics of MetS and control groups in Clusters 4–6 (year −1 to 0).** Categorical variables are expressed as number (%). For each variable, participants with missing values were excluded from the variable-specific denominators. The group-specific sample sizes shown in the row labeled “n” represent the total number of participants in each group, including those with missing lifestyle data.

**S1 Fig. Encoding process of MetS components for clustering analysis.** Abbreviations: MetS, metabolic syndrome; WC, waist circumference (three-level classification); HT, hypertension (three-level classification); DLP, dyslipidemia (three-level classification); HG, hyperglycemia (three-level classification).

**S2 Fig. Silhouette scores and ARI for cluster number evaluation and method comparison.** Abbreviations: ARI, adjusted Rand index.

**S3 Fig. Heatmaps showing the distribution of MetS component combinations across Clusters 1–6 in the MetS group.** Clusters 1–3 (top) and Clusters 4–6 (bottom) are displayed vertically. Each row represents a unique combination of MetS diagnostic components, including OB, HT, DLP, and HG. “None” indicates individuals who met none of the diagnostic criteria for any of the four components. Each column corresponds to a specific year within each cluster, from year −3 to 0 (left to right). Cell values indicate the proportion of individuals with each component combination, calculated as a percentage of the total number of individuals in that column. The percentage is annotated within each cell. Color intensity reflects the proportion, ranging from 0% (lightest) to 100% (darkest). Cells with no individuals are marked with an en dash (–). Abbreviations: MetS, metabolic syndrome; OB, abdominal obesity; HT, hypertension; DLP, dyslipidemia; HG, hyperglycemia.

**S4 Fig. Heatmaps showing the distribution of MetS component combinations across Clusters 1–6 in the control group.** Clusters 1–3 (top) and Clusters 4–6 (bottom) are displayed vertically. Each row represents a unique combination of MetS diagnostic components, including OB, HT, DLP, and HG. “None” indicates individuals who met none of the diagnostic criteria for any of the four components. Each column corresponds to a specific year within each cluster, from year −3 to 0 (left to right). Cell values indicate the proportion of individuals with each component combination, calculated as a percentage of the total number of individuals in that column. The percentage is annotated within each cell. Color intensity reflects the proportion, ranging from 0% (lightest) to 100% (darkest). Cells with no individuals are marked with an en dash (–). Abbreviations: OB, abdominal obesity; HT, hypertension; DLP, dyslipidemia; HG, hyperglycemia.

**S5 Fig. Cluster-specific WC change.** Abbreviations: MetS, metabolic syndrome; WC, waist circumference.

**S6 Fig. Predictors of annual WC change from year −3 to −1.** Results are based on multiple linear regression analyses examining factors associated with annual changes in WC. The top panel shows results for the combined group (MetS and control), and the bottom panel shows results for the MetS group alone. Definitions and descriptions of the lifestyle variables are provided in S1–S3 Tables. Abbreviations: MetS, metabolic syndrome; WC, waist circumference.

**S7 Fig. Predictors of annual WC change from year −1 to 0.** Results are based on multiple linear regression analyses examining factors associated with annual changes in WC. The top panel shows results for the combined group (MetS and control), and the bottom panel shows results for the MetS group alone. Definitions and descriptions of the lifestyle variables are provided in S1–S3 Tables. Abbreviations: MetS, metabolic syndrome; WC, waist circumference.

S8 Fig. Diagnosis-positive rates for each MetS component—OB, HT, DLP, and HG—in females.

Females included in the analysis were n = 56 (median age: 55.0 years; interquartile range: 54.0–57.0), with age values corresponding to year −3.

Abbreviations: MetS, metabolic syndrome; OB, abdominal obesity; HT, hypertension; DLP, dyslipidemia; HG, hyperglycemia.

Abbreviations: MetS, metabolic syndrome; OB, abdominal obesity; HT, hypertension; DLP, dyslipidemia; HG, hyperglycemia.

**S9 Fig. Diagnosis-positive rates for each MetS component—OB, HT, DLP, and HG—in males, with analyses covering the period from year −5 to 0.**

Males included in the analysis were n = 139 (median age: 54.0 years; interquartile range: 52.0–55.0), with age values corresponding to year −5.

Abbreviations: MetS, metabolic syndrome; OB, abdominal obesity; HT, hypertension; DLP, dyslipidemia; HG, hyperglycemia.

Abbreviations: MetS, metabolic syndrome; OB, abdominal obesity; HT, hypertension; DLP, dyslipidemia; HG, hyperglycemia.

## Notes

### Competing Interest Statement

The authors have declared no competing interest.

### Author Declarations

Ethical approval was granted by the Ethics Committee of Toyama University Hospital (approval number R2021070; approval date 19 August 2021). All participants provided written informed consent.

## References

1. Ford ES. Risks for all-cause mortality, cardiovascular disease, and diabetes associated with the metabolic syndrome: a summary of the evidence. Diabetes Care. 2005;28(7):1769–1778. doi: 10.2337/diacare.28.7.1769.

2. Després JP, Lemieux I. Abdominal obesity and metabolic syndrome. Nature. 2006;444(7121):881–887. doi: 10.1038/nature05488.

3. Neeland IJ, Lim S, Tchernof A, Gastaldelli A, Rangaswami J, Ndumele CE, et al. Metabolic syndrome. Nat Rev Dis Primers. 2024;10(1):77. doi: 10.1038/s41572-024-00563-5.

4. Loftus TJ, Shickel B, Balch JA, Tighe PJ, Abbott KL, Fazzone B, et al. Phenotype clustering in health care: A narrative review for clinicians. Front Artif Intell. 2022;5:842306. doi: 10.3389/frai.2022.842306.

5. Shikata M, Oku M, Fukuhara S, Ito R, Haruki T, Ueda K, et al. Characterization of individuals in whom body weight loss precedes diabetes onset: a retrospective, observational, longitudinal cohort study based on health checkup in Japan. Endocr J. 2026;73(1):43–52. doi: 10.1507/endocrj.EJ25-0230.

6. Ito R, Oku M, Kimura I, Haruki T, Shikata M, Teramoto T, et al. Energy landscape analysis of health checkup data clarified multiple pathways to diabetes development in obese and non-obese subjects. Front Endocrinol (Lausanne). 2025;16:1576431. doi: 10.3389/fendo.2025.1576431.

7. Lim AMW, Lim EU, Chen PL, Fann CSJ. Unsupervised clustering identified clinically relevant metabolic syndrome endotypes in UK and Taiwan Biobanks. iScience. 2024;27(7):109815. doi: 10.1016/j.isci.2024.109815.

8. Kim J. Metabotype risk clustering based on metabolic disease biomarkers and its association with metabolic syndrome in Korean adults: findings from the 2016–2023 Korea National Health and Nutrition Examination Survey (KNHANES). Diseases. 2025;13(8):239. doi: 10.3390/diseases13080239.

9. Park SJ, Kim YN, Oh BK, Kang J. Risk factors for metabolic syndrome in the premetabolic state assessed using hierarchical clustering study in a health screening group. Sci Rep. 2024;14(1):31169. doi: 10.1038/s41598-024-82513-5.

10. Riedl A, Wawro N, Gieger C, Meisinger C, Peters A, Roden M, et al. Identification of comprehensive metabotypes associated with cardiometabolic diseases in the population-based KORA study. Mol Nutr Food Res. 2018;62(16):e1800117. doi: 10.1002/mnfr.201800117.

11. Beck JR, Pauker SG. The Markov process in medical prognosis. Med Decis Making. 1983;3(4):419–458. doi: 10.1177/0272989X8300300403.

12. Sonnenberg FA, Beck JR. Markov models in medical decision making: a practical guide. Med Decis Making. 1993;13(4):322–338. doi: 10.1177/0272989X9301300409.

13. Hwang LC, Bai CH, You SL, Sun CA, Chen CJ. Description and prediction of the development of metabolic syndrome: a longitudinal analysis using a Markov model approach. PLoS One. 2013;8(6):e67436. doi: 10.1371/journal.pone.0067436.

14. Chen X, Chen Q, Chen L, Zhang P, Xiao J, Wang S. Description and prediction of the development of metabolic syndrome in Dongying City: a longitudinal analysis using the Markov model. BMC Public Health. 2014;14:1033. doi: 10.1186/1471-2458-14-1033.

15. Jia X, Chen Q, Wu P, Liu M, Chen X, Xiao J, et al. Dynamic development of metabolic syndrome and its risk prediction in Chinese population: a longitudinal study using Markov model. Diabetol Metab Syndr. 2018;10:24. doi: 10.1186/s13098-018-0328-3.

16. Bagheri P, Khalili D, Seif M, Rezaianzadeh A. Dynamic behavior of metabolic syndrome progression: a comprehensive systematic review on recent discoveries. BMC Endocr Disord. 2021;21(1):54. doi: 10.1186/s12902-021-00716-7.

17. Matsuzawa Y. Metabolic syndrome--definition and diagnostic criteria in Japan. J Atheroscler Thromb. 2005;12(6):301. doi: 10.5551/jat.12.301.

18. Sacks DB, Arnold M, Bakris GL, Bruns DE, Horvath AR, Lernmark Å, et al. Guidelines and recommendations for laboratory analysis in the diagnosis and management of diabetes mellitus. Diabetes Care. 2023;46(10):e151–e199. doi: 10.2337/dci23-0036.

19. Ministry of Health, Labour and Welfare. Standard program for specific health checkups and guidance [Internet]. Tokyo: Ministry of Health, Labour and Welfare; 2024 [cited 2026 Mar 1]. Available from: https://www.mhlw.go.jp/content/10900000/001231390.pdf

20. Tsushita K, S Hosler A, Miura K, Ito Y, Fukuda T, Kitamura A, et al. Rationale and descriptive analysis of specific health guidance: the nationwide lifestyle intervention program targeting metabolic syndrome in Japan. J Atheroscler Thromb. 2018;25(4):308–322. doi: 10.5551/jat.42010.

21. Jonker R, Volgenant A. A shortest augmenting path algorithm for dense and sparse linear assignment problems. Computing. 1987;38:325–340. doi: 10.1007/BF02278710.

22. Murtagh F, Contreras P. Algorithms for hierarchical clustering: an overview. WIREs Data Min Knowl Discov. 2012;2:86–97. doi: 10.1002/widm.53.

23. Chacón JE, Rastrojo AI. Minimum adjusted Rand index for two clusterings of a given size. Adv Data Anal Classif. 2023;17:125–133. doi: 10.1007/s11634-022-00491-w.

24. Franco OH, Massaro JM, Civil J, Cobain MR, O’Malley B, D’Agostino RB Sr. Trajectories of entering the metabolic syndrome: the framingham heart study. Circulation. 2009;120(20):1943–1950. doi: 10.1161/CIRCULATIONAHA.109.855817.

25. Rao X, Xin Z, Yu Q, Feng L, Shi Y, Tang T, et al. Triglyceride-glucose-body mass index and the incidence of cardiovascular diseases: a meta-analysis of cohort studies. Cardiovasc Diabetol. 2025;24(1):34. doi: 10.1186/s12933-025-02584-0.

26. Dang K, Wang X, Hu J, Zhang Y, Cheng L, Qi X, et al. The association between triglyceride-glucose index and its combination with obesity indicators and cardiovascular disease: NHANES 2003-2018. Cardiovasc Diabetol. 2024;23(1):8. doi: 10.1186/s12933-023-02115-9.

27. Nayak SS, Kuriyakose D, Polisetty LD, Patil AA, Ameen D, Bonu R, et al. Diagnostic and prognostic value of triglyceride glucose index: a comprehensive evaluation of meta-analysis. Cardiovasc Diabetol. 2024;23(1):310. doi: 10.1186/s12933-024-02392-y.

28. Lee J, Kim B, Kim W, Ahn C, Choi HY, Kim JG, et al. Lipid indices as simple and clinically useful surrogate markers for insulin resistance in the U.S. population. Sci Rep. 2021;11(1):2366. doi: 10.1038/s41598-021-82053-2.

29. Kramer CK, Zinman B, Retnakaran R. Are metabolically healthy overweight and obesity benign conditions?: A systematic review and meta-analysis. Ann Intern Med. 2013;159(11):758–769. doi: 10.7326/0003-4819-159-11-201312030-00008.

30. Hiramatsu Y, Ide H, Furui Y. Differences in the components of metabolic syndrome by age and sex: a cross-sectional and longitudinal analysis of a cohort of middle-aged and older Japanese adults. BMC Geriatr. 2023;23(1):438. doi: 10.1186/s12877-023-04145-0.

31. Pucci G, Alcidi R, Tap L, Battista F, Mattace-Raso F, Schillaci G. Sex- and gender-related prevalence, cardiovascular risk and therapeutic approach in metabolic syndrome: A review of the literature. Pharmacol Res. 2017;120:34–42. doi: 10.1016/j.phrs.2017.03.008.

32. Ge H, Yang Z, Li X, Liu D, Li Y, Pan Y, et al. The prevalence and associated factors of metabolic syndrome in Chinese aging population. Sci Rep. 2020;10(1):20034. doi: 10.1038/s41598-020-77184-x.

33. Rodriguez A, Muller DC, Metter EJ, Maggio M, Harman SM, Blackman MR, et al. Aging, androgens, and the metabolic syndrome in a longitudinal study of aging. J Clin Endocrinol Metab. 2007;92(9):3568–3572. doi: 10.1210/jc.2006-2764.

34. Kuzuya M, Ando F, Iguchi A, Shimokata H. Age-specific change of prevalence of metabolic syndrome: longitudinal observation of large Japanese cohort. Atherosclerosis. 2007;191(2):305–312. doi: 10.1016/j.atherosclerosis.2006.05.043.

35. Cho JH, Kwon HM, Park SE, Jung JH, Han KD, Park YG, et al. Protective effect of smoking cessation on subsequent myocardial infarction and ischemic stroke independent of weight gain: A nationwide cohort study. PLoS One. 2020;15(7):e0235276. doi: 10.1371/journal.pone.0235276.

36. Kos K. Cardiometabolic morbidity and mortality with smoking cessation, review of recommendations for people with diabetes and obesity. Curr Diab Rep. 2020;20(12):82. doi: 10.1007/s11892-020-01352-6.

37. Deng Y, Yang Q, Hao C, Wang HH, Ma T, Chen X, et al. Combined lifestyle factors and metabolic syndrome risk: a systematic review and meta-analysis. Int J Obes (Lond). 2025;49(2):226–236. doi: 10.1038/s41366-024-01671-8.

38. Islam MS, Wei P, Suzauddula M, Nime I, Feroz F, Acharjee M, et al. The interplay of factors in metabolic syndrome: understanding its roots and complexity. Mol Med. 2024;30(1):279. doi: 10.1186/s10020-024-01019-y.

39. Razbek J, Zhang Y, Xia WJ, Xu WT, Li DY, Yin Z, et al. Study on dynamic progression and risk assessment of metabolic syndrome based on multi-state Markov model. Diabetes Metab Syndr Obes. 2022;15:2497–2510. doi: 10.2147/DMSO.S362071.

40. Gao CX, Dwyer D, Zhu Y, Smith CL, Du L, Filia KM, et al. An overview of clustering methods with guidelines for application in mental health research. Psychiatry Res. 2023;327:115265. doi: 10.1016/j.psychres.2023.115265.

