## Supplementary_Information for "Multiple pathways to metabolic syndrome identified by hierarchical clustering of Japanese longitudinal health checkup data"

Title

**S1 Table. Binary coding of questionnaire responses.**

This table provides binary coding definitions for self-administered questionnaire items used in lifestyle characteristics (S11–S14 Tables) and multivariable regression analyses.

| Questionnaire Item | Binary Value | Definition |
|---|---|---|
| Current smoking “Do you smoke cigarettes regularly?” | 0 | Never smoked or formerly smoked |
| | 1 | Currently smoke |
| Frequency of drinking “How often do you drink alcohol?” | 0 | Sometimes or rarely/never |
| | 1 | Every day |
| Lifestyle efforts “Do you intend to improve your lifestyle habits, such as exercise and diet?” | 0 | No intention to improve or not yet actively improving |
| | 1 | Improving for 6 months or more |
| Willingness to receive guidance “Would you be willing to receive health guidance?” | 0 | No |
| | 1 | Yes |

**S2 Table. Variable definitions (year −3 to −1).**

Variables derived from questionnaire responses (S1 Table), collected during the period from year −3 to −1.

| Variable | Definition | Reference Category |
|---|---|---|
| Frequency of smoking | Proportion of respondents who answered 1 to *Current smoking* (range: 0.0–1.0) | – |
| Smoking cessation | Changed response from 1 to 0 for *Current smoking* | No change |
| Frequency of daily drinking | Proportion of respondents who answered 1 to *Frequency of drinking* (range: 0.0–1.0) | – |
| Frequency of lifestyle efforts | Proportion of respondents who answered 1 to *Lifestyle efforts* (range: 0.0–1.0) | – |
| Willingness to receive guidance | Answered 1 to *Willingness to receive guidance* during year −3 to −1 | Never willing |

**S3 Table. Variable definitions (year −1 to 0).**

Variables derived from questionnaire responses (S1 Table), collected during the period from year −1 to 0.

| Variable | Definition | Reference Category |
|---|---|---|
| Smoking cessation | Change in response from 1 to 0 for *Current smoking* | No change |
| Shift toward daily drinking | Change in response from 0 to 1 for *Frequency of drinking* | No change |
| Ceased daily drinking | Change in response from 1 to 0 for *Frequency of drinking* | No change |
| Reduced lifestyle effort | Change in response from 1 to 0 for *Lifestyle efforts* | Maintained effort |
| Willingness to receive guidance | Answered 1 to *Willingness to receive guidance* during year −3 to 0 | Never willing |

**S4 Table. Criteria for three-level classification of MetS components.**

| Component | Score 0 (Normal) | Score 1 (Intermediate) | Score 2 (MetS Criteria) |
|---|---|---|---|
| Abdominal Obesity | Male: WC < 80 cm | Male: 80 cm ≤ WC < 85 cm | Male: WC ≥ 85 cm |
| | Female: WC < 85 cm | Female: 85 cm ≤ WC < 90 cm | Female: WC ≥ 90 cm |
| Hypertension | SBP < 120 mmHg AND DBP < 80 mmHg AND no antihypertensive medication | (SBP ≥ 120 mmHg OR DBP ≥ 80 mmHg) AND (SBP < 130 mmHg AND DBP < 85 mmHg) AND no antihypertensive medication | SBP ≥ 130 mmHg OR DBP ≥ 85 mmHg OR receiving antihypertensive medication |
| Dyslipidemia | TG < 100 mg/dL AND HDL-C ≥ 60 mg/dL AND no lipid-lowering medication | (TG ≥ 100 mg/dL OR HDL-C < 60 mg/dL) AND TG < 150 mg/dL AND HDL-C ≥ 40 mg/dL AND no lipid-lowering medication | TG ≥ 150 mg/dL OR HDL-C < 40 mg/dL OR receiving lipid-lowering medication |
| Hyperglycemia | FPG < 100 mg/dL AND HbA1c < 5.6% AND no antidiabetic medication AND no diagnosis with diabetes | (FPG ≥ 100 mg/dL OR HbA1c ≥ 5.6%) AND FPG < 110 mg/dL AND HbA1c < 6.0% AND no antidiabetic medication AND no diagnosis with diabetes | FPG ≥ 110 mg/dL OR HbA1c ≥ 6.0% OR receiving antidiabetic medication OR diagnosis with diabetes |

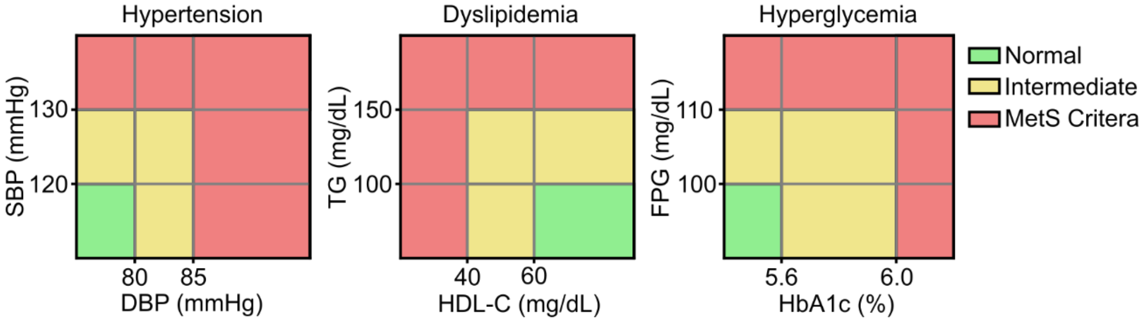


Abbreviations: MetS, metabolic syndrome; WC, waist circumference; SBP, systolic blood pressure; DBP, diastolic blood pressure; TG, triglycerides; HDL-C, high-density lipoprotein cholesterol; FPG, fasting plasma glucose; HbA1c, glycated hemoglobin.

**S5 Table. Background characteristics of study participants (year −3).**

| | MetS | Control | P-value |
|---|---|---|---|
| n | 296 | 296 | — |
| Age [years] | 55.0 [53.0–56.0] | 56.0 [54.0–58.0] | <0.001 |
| Age ≥ 55 years, n (%) | 156 (52.7) | 218 (73.6) | <0.001 |
| WC [cm] | 85.6 [82.5–90.2] | 85.3 [82.5–88.7] | 0.313 |
| WC ≥ 85 cm, n (%) | 160 (54.1) | 159 (53.7) | 1.000 |
| BMI [kg/m$^2$] | 23.8 [22.3–25.3] | 23.5 [22.4–25.0] | 0.278 |
| BMI ≥ 25, n (%) | 87 (29.4) | 75 (25.3) | 0.311 |
| SBP [mmHg] | 127.0 [119.0–134.0] | 128.0 [119.0–135.0] | 0.784 |
| DBP [mmHg] | 80.0 [73.0–86.0] | 80.5 [72.0–86.0] | 0.914 |
| HT, n (%) | 160 (54.1) | 160 (54.1) | 1.0 |
| TG [mg/dL] | 111.0 [79.8–148.2] | 108.5 [80.0–146.2] | 0.620 |
| HDL-C [mg/dL] | 55.0 [46.8–64.0] | 56.0 [49.8–66.0] | 0.068 |
| DLP, n (%) | 91 (30.7) | 89 (30.1) | 0.929 |
| PG [mg/dL] | 94.0 [88.0–100.0] | 93.0 [88.0–100.0] | 0.640 |
| HbA1c [%] | 5.7 [5.5–5.8] | 5.6 [5.4–5.8] | 0.124 |
| HG, n (%) | 39 (13.2) | 39 (13.2) | 1.0 |
| Diabetes, n (%) | 11 (3.7) | 11 (3.7) | 1.0 |
| MetS component, n (%) | — | — | 0.998 |
| 0 | 57 (19.3) | 59 (19.9) | — |
| 1 | 195 (65.9) | 193 (65.2) | — |
| 2 | 37 (12.5) | 37 (12.5) | — |
| 3 | 7 (2.4) | 7 (2.4) | — |
| TyG-WC | 740.2 [704.4–787.7] | 737.0 [699.3–768.1] | 0.087 |
| TyG-BMI | 203.8 [191.3–219.3] | 201.8 [188.7–217.8] | 0.111 |

All continuous variables were non-normally distributed and are presented as median [interquartile range]. Categorical variables are expressed as number (%).

“MetS component, n (%)” indicates the number of components meeting the diagnostic criteria for MetS among the following: HT, DLP, HG, excluding OB.
Abbreviations: MetS, metabolic syndrome; WC, waist circumference; BMI, body mass index; SBP, systolic blood pressure; DBP, diastolic blood pressure; HT, hypertension; TG, triglycerides; HDL-C, high-density lipoprotein cholesterol; DLP, dyslipidemia; PG, plasma glucose; HbA1c, glycated hemoglobin; HG, hyperglycemia; TyG-WC, triglyceride-glucose index × waist circumference; TyG-BMI, triglyceride-glucose index × body mass index.

**S6 Table. Clinical characteristics of MetS and control groups at year −3 (Clusters 1–3).**

| | Cluster1 MetS | Cluster1 Control | Cluster1 p-value | Cluster2 MetS | Cluster2 Control | Cluster2 p-value | Cluster3 MetS | Cluster3 Control | Cluster3 p-value |
|---|---|---|---|---|---|---|---|---|---|
| n | 97 | 97 | — | 36 | 36 | — | 48 | 48 | — |
| Age [years] | 55.0 [52.0–57.0] | 56.0 [54.0–57.0] | 0.008 | 54.0 [52.0–56.0] | 56.0 [54.5–57.0] | 0.048 | 54.5 ± 2.5 | 56.4 ± 2.2 | < 0.001 |
| Age ≥ 55 years, n (%) | 52 (53.6) | 64 (66.0) | 0.107 | 16 (44.4) | 27 (75.0) | 0.016 | 22 (45.8) | 43 (89.6) | < 0.001 |
| WC [cm] | 88.9 [85.7–94.2] | 87.2 [85.2–90.8] | 0.093 | 87.3 [85.8–93.1] | 88.5 [86.0–92.4] | 0.946 | 89.0 [87.1–93.2] | 88.0 [86.7–91.5] | 0.149 |
| WC ≥ 85 cm, n (%) | 79 (81.4) | 79 (81.4) | 1.0 | 30 (83.3) | 30 (83.3) | 1.0 | 47 (97.9) | 46 (95.8) | 1.000 |
| BMI [kg/m$^2$] | 24.9 [23.5–26.4] | 24.4 [23.3–26.0] | 0.287 | 24.1 [23.0–26.2] | 24.4 [23.6–25.7] | 0.660 | 24.8 [23.8–26.2] | 24.4 [22.8–25.8] | 0.181 |
| BMI ≥ 25, n (%) | 46 (47.4) | 41 (42.3) | 0.564 | 13 (36.1) | 11 (30.6) | 0.803 | 23 (47.9) | 18 (37.5) | 0.409 |
| SBP [mmHg] | 133.0 [127.0–144.0] | 132.0 [127.0–138.0] | 0.139 | 114.9 ± 8.6 | 115.7 ± 8.2 | 0.696 | 123.0 [117.0–127.2] | 122.5 [112.0–128.0] | 0.655 |
| DBP [mmHg] | 86.0 [80.0–92.0] | 84.0 [77.0–88.0] | 0.064 | 70.8 ± 5.7 | 71.8 ± 5.5 | 0.464 | 76.0 [71.0–80.0] | 76.0 [69.0–81.0] | 0.918 |
| HT, n (%) | 86 (88.7) | 86 (88.7) | 1.0 | 1 (2.8) | 1 (2.8) | 1.0 | 7 (14.6) | 7 (14.6) | 1.0 |
| TG [mg/dL] | 94.0 [73.0–119.0] | 93.0 [69.0–118.0] | 0.582 | 145.5 [119.5–268.8] | 143.0 [106.5–169.2] | 0.148 | 103.5 [71.0–126.5] | 103.5 [78.5–131.0] | 0.769 |
| HDL-C [mg/dL] | 58.0 [52.0–66.0] | 59.0 [53.0–70.0] | 0.374 | 46.0 [40.8–50.5] | 50.5 [45.5–57.5] | 0.024 | 53.0 [47.8–62.0] | 53.0 [46.8–61.0] | 0.915 |
| DLP, n (%) | 4 (4.1) | 4 (4.1) | 1.0 | 25 (69.4) | 23 (63.9) | 0.803 | 6 (12.5) | 6 (12.5) | 1.0 |
| PG [mg/dL] | 92.4 ± 6.8 | 91.7 ± 7.0 | 0.526 | 92.6 ± 6.8 | 91.4 ± 7.3 | 0.495 | 96.4 ± 9.7 | 96.9 ± 9.4 | 0.797 |
| HbA1c [%] | 5.5 [5.3–5.7] | 5.5 [5.3–5.6] | 0.161 | 5.7 [5.5–5.8] | 5.6 [5.4–5.8] | 0.941 | 5.7 [5.7–5.9] | 5.7 [5.6–5.9] | 0.159 |
| HG, n (%) | 0 (0.0) | 0 (0.0) | — | 1 (2.8) | 1 (2.8) | 1.0 | 12 (25.0) | 12 (25.0) | 1.0 |
| Diabetes, n (%) | 0 (0.0) | 0 (0.0) | — | 0 (0.0) | 0 (0.0) | — | 3 (6.2) | 0 (0.0) | 0.242 |
| Mets component, n (%) | — | — | 1.0 | — | — | 0.792 | — | — | 1.0 |
| 0 | 8 (8.2) | 8 (8.2) | — | 9 (25.0) | 11 (30.6) | — | 23 (47.9) | 23 (47.9) | — |
| 1 | 88 (90.7) | 88 (90.7) | — | 27 (75.0) | 25 (69.4) | — | 25 (52.1) | 25 (52.1) | — |
| 2 | 1 (1.0) | 1 (1.0) | — | 0 (0.0) | 0 (0.0) | — | 0 (0.0) | 0 (0.0) | — |
| 3 | 0 (0.0) | 0 (0.0) | — | 0 (0.0) | 0 (0.0) | — | 0 (0.0) | 0 (0.0) | — |
| TyG-WC | 747.5 [708.9–794.7] | 728.9 [704.9–765.0] | 0.102 | 802.6 ± 71.5 | 780.9 ± 54.3 | 0.151 | 764.8 ± 44.2 | 759.7 ± 54.9 | 0.619 |
| TyG-BMI | 208.5 [195.4–222.6] | 202.8 [192.5–217.8] | 0.145 | 221.3 ± 26.3 | 217.0 ± 20.7 | 0.448 | 212.0 ± 16.0 | 209.0 ± 21.6 | 0.440 |

Values are presented as mean ± SD or median [interquartile range], depending on data distribution. Categorical variables are expressed as number (%). “MetS component, n (%)” indicates the number of components meeting the diagnostic criteria for MetS among the following: HT, DLP, HG, excluding OB. Abbreviations: MetS, metabolic syndrome; WC, waist circumference; BMI, body mass index; SBP, systolic blood pressure; DBP, diastolic blood pressure; HT, hypertension; TG, triglycerides; HDL-C, high-density lipoprotein cholesterol; DLP, dyslipidemia; PG, plasma glucose; HbA1c, glycated hemoglobin; HG, hyper glycemia; TyG-WC, triglyceride-glucose index × waist circumference; TyG-BMI, triglyceride-glucose index × body mass index.

**S7 Table. Clinical characteristics of MetS and control groups at year −3 (Clusters 4–6).**

| | Cluster4 MetS | Cluster4 Control | Cluster4 p-value | Cluster5 MetS | Cluster5 Control | Cluster5 p-value | Cluster6 MetS | Cluster6 Control | Cluster6 p-value |
|---|---|---|---|---|---|---|---|---|---|
| n | 44 | 44 | — | 46 | 46 | — | 25 | 25 | — |
| Age [years] | 54.4 ± 2.5 | 55.3 ± 3.0 | 0.131 | 55.0 [53.2–57.0] | 56.0 [54.0–58.0] | 0.121 | 55.7 ± 2.1 | 57.0 ± 2.4 | 0.042 |
| Age ≥ 55 years, n (%) | 22 (50.0) | 30 (68.2) | 0.129 | 27 (58.7) | 31 (67.4) | 0.517 | 17 (68.0) | 23 (92.0) | 0.077 |
| WC [cm] | 81.8 [79.8–83.4] | 81.8 [79.8–83.2] | 0.748 | 81.7 ± 3.0 | 81.6 ± 2.9 | 0.916 | 82.5 [81.0–83.0] | 82.6 [81.2–83.5] | 0.838 |
| WC ≥ 85 cm, n (%) | 1 (2.3) | 1 (2.3) | 1.0 | 3 (6.5) | 3 (6.5) | 1.0 | 0 (0.0) | 0 (0.0) | — |
| BMI [kg/m$^2$] | 22.5 [21.5–23.3] | 22.8 [21.6–23.3] | 0.881 | 22.6 ± 1.4 | 22.6 ± 1.7 | 0.871 | 22.7 ± 1.7 | 21.7 ± 1.3 | 0.026 |
| BMI ≥ 25, n (%) | 0 (0.0) | 1 (2.3) | 1.000 | 3 (6.5) | 4 (8.7) | 1.000 | 2 (8.0) | 0 (0.0) | 0.490 |
| SBP [mmHg] | 128.3 ± 11.9 | 129.0 ± 10.7 | 0.764 | 128.0 [122.5–132.8] | 129.0 [125.0–139.2] | 0.158 | 122.2 ± 13.7 | 125.8 ± 11.6 | 0.323 |
| DBP [mmHg] | 79.4 ± 9.1 | 81.9 ± 8.9 | 0.197 | 81.0 ± 6.9 | 82.2 ± 8.5 | 0.471 | 76.6 ± 9.6 | 78.7 ± 10.0 | 0.458 |
| HT, n (%) | 26 (59.1) | 26 (59.1) | 1.0 | 27 (58.7) | 27 (58.7) | 1.0 | 13 (52.0) | 13 (52.0) | 1.0 |
| TG [mg/dL] | 187.0 [153.8–226.8] | 170.0 [154.0–211.5] | 0.652 | 86.5 [69.2–111.8] | 84.0 [68.0–111.8] | 0.953 | 155.0 [107.0–205.0] | 133.0 [106.0–200.0] | 0.861 |
| HDL-C [mg/dL] | 52.0 [44.0–64.2] | 56.0 [49.2–64.0] | 0.196 | 62.0 [51.0–69.8] | 63.0 [53.0–69.8] | 0.639 | 54.6 ± 12.3 | 55.3 ± 15.0 | 0.861 |
| DLP, n (%) | 39 (88.6) | 39 (88.6) | 1.0 | 0 (0.0) | 0 (0.0) | — | 17 (68.0) | 17 (68.0) | 1.0 |
| PG [mg/dL] | 92.7 ± 7.2 | 92.1 ± 7.0 | 0.697 | 96.5 [90.0–103.5] | 96.0 [91.2–102.8] | 0.611 | 106.0 [96.0–118.0] | 107.0 [97.0–116.0] | 1.000 |
| HbA1c [%] | 5.5 [5.4–5.7] | 5.5 [5.4–5.7] | 0.654 | 5.7 [5.5–5.8] | 5.7 [5.5–5.8] | 0.924 | 6.0 [5.9–6.3] | 6.0 [5.6–6.2] | 0.647 |
| HG, n (%) | 1 (2.3) | 1 (2.3) | 1.0 | 6 (13.0) | 6 (13.0) | 1.0 | 19 (76.0) | 19 (76.0) | 1.0 |
| Diabetes, n (%) | 0 (0.0) | 0 (0.0) | — | 1 (2.2) | 2 (4.3) | 1.000 | 7 (28.0) | 9 (36.0) | 0.762 |
| MetS component, n (%) | — | — | 1.0 | — | — | 1.0 | — | — | 1.0 |
| 0 | 1 (2.3) | 1 (2.3) | — | 16 (34.8) | 16 (34.8) | — | 0 (0.0) | 0 (0.0) | — |
| 1 | 20 (45.5) | 20 (45.5) | — | 27 (58.7) | 27 (58.7) | — | 8 (32.0) | 8 (32.0) | — |
| 2 | 23 (52.3) | 23 (52.3) | — | 3 (6.5) | 3 (6.5) | — | 10 (40.0) | 10 (40.0) | — |
| 3 | 0 (0.0) | 0 (0.0) | — | 0 (0.0) | 0 (0.0) | — | 7 (28.0) | 7 (28.0) | — |
| TyG-WC | 743.2 [708.4–768.2] | 738.6 [706.7–752.4] | 0.416 | 680.3 ± 43.6 | 681.2 ± 39.5 | 0.923 | 742.5 ± 57.1 | 745.8 ± 68.6 | 0.855 |
| TyG-BMI | 202.6 ± 20.1 | 200.7 ± 17.1 | 0.645 | 187.8 ± 14.5 | 188.7 ± 17.1 | 0.782 | 199.5 [186.2–223.0] | 199.3 [187.1–204.3] | 0.383 |

Values are presented as mean ± SD or median [interquartile range], depending on data distribution. Categorical variables are expressed as number (%). “MetS component, n (%)” indicates the number of components meeting the diagnostic criteria for MetS among the following: HT, DLP, HG, excluding OB. Abbreviations: MetS, metabolic syndrome; WC, waist circumference; BMI, body mass index; SBP, systolic blood pressure; DBP, diastolic blood pressure; HT, hypertension; TG, triglycerides; HDL-C, high-density lipoprotein cholesterol; DLP, dyslipidemia; PG, plasma glucose; HbA1c, glycated hemoglobin; HG, hyper glycemia; TyG-WC, triglyceride-glucose index × waist circumference; TyG-BMI, triglyceride-glucose index × body mass index.

**S8 Table. Detailed definitions and descriptions of selected variables.**

| | Year −3 | Year −2 | Year −1 | Year 0 |
|---|---|---|---|---|
| Decreasing trend (Year −3 to −1) | Yes | No | No | — |
| | Yes | Yes | No | — |
| Unchanging trend (Year −3 to −1) | No | No | No | — |
| | Yes | Yes | Yes | — |
| Temporary shift (Year −3 to −1) | Yes | No | Yes | — |
| | No | Yes | No | — |
| Increasing trend (Year −3 to −1) | No | No | Yes | — |
| | No | Yes | Yes | — |
| Decreasing change (Year −1 to 0) | — | — | Yes | No |
| No change (Year −1 to 0) | — | — | No | No |
| | — | — | Yes | Yes |
| Increasing change (Year −1 to 0) | — | — | No | Yes |

“Yes” indicates that the individual met the diagnostic criterion for the corresponding MetS component (HT, DLP, or HG) in that year. “No” indicates that the criterion was not met.

Abbreviations: MetS, metabolic syndrome; HT, hypertension; DLP, dyslipidemia; HG, hyper glycemia.

**S9 Table. Clinical characteristics of MetS and control groups at year 0 in Clusters 1–3.**

| | Cluster1 MetS | Cluster1 Control | Cluster1 p-value | Cluster2 MetS | Cluster2 Control | Cluster2 p-value | Cluster3 MetS | Cluster3 Control | Cluster3 p-value |
|---|---|---|---|---|---|---|---|---|---|
| n | 97 | 97 | — | 36 | 36 | — | 48 | 48 | — |
| WC [cm] | 90.0 [88.0–95.6] | 86.2 [83.9–90.5] | < 0.001 | 92.3 ± 5.8 | 88.0 ± 6.4 | 0.004 | 92.0 ± 4.7 | 88.3 ± 4.7 | < 0.001 |
| WC ≥ 85 cm, n (%) | 97 (100.0) | 59 (60.8) | < 0.001 | 36 (100.0) | 22 (61.1) | < 0.001 | 48 (100.0) | 36 (75.0) | < 0.001 |
| BMI [kg/m$^2$] | 25.4 [24.2–26.9] | 24.1 [22.9–25.8] | < 0.001 | 25.0 [24.1–26.1] | 24.0 [22.9–25.6] | 0.121 | 25.7 ± 2.1 | 24.3 ± 2.1 | 0.003 |
| BMI ≥ 25, n (%) | 58 (59.8) | 37 (38.1) | 0.004 | 19 (52.8) | 13 (36.1) | 0.236 | 28 (58.3) | 16 (33.3) | 0.024 |
| SBP [mmHg] | 136.0 [129.0–150.0] | 133.0 [127.0–140.0] | 0.011 | 127.7 ± 8.5 | 118.0 ± 9.0 | < 0.001 | 132.0 [130.0–136.2] | 123.5 [114.8–135.0] | 0.001 |
| DBP [mmHg] | 88.0 [82.0–94.0] | 83.0 [78.0–92.0] | 0.008 | 79.6 ± 7.2 | 75.0 ± 6.8 | 0.007 | 84.0 [80.0–87.0] | 78.0 [71.8–84.2] | 0.001 |
| HT, n (%) | 96 (99.0) | 75 (77.3) | < 0.001 | 25 (69.4) | 3 (8.3) | < 0.001 | 44 (91.7) | 18 (37.5) | < 0.001 |
| TG [mg/dL] | 156.0 [88.0–173.0] | 87.0 [65.0–112.0] | < 0.001 | 192.5 [131.0–278.8] | 141.0 [88.0–174.8] | 0.003 | 115.5 [89.0–161.2] | 83.0 [65.5–125.0] | 0.005 |
| HDL-C [mg/dL] | 58.0 [49.0–65.0] | 58.0 [52.0–73.0] | 0.032 | 46.5 ± 9.2 | 52.8 ± 8.9 | 0.004 | 53.0 [47.0–66.0] | 54.5 [49.0–64.2] | 0.519 |
| DLP, n (%) | 71 (73.2) | 7 (7.2) | < 0.001 | 35 (97.2) | 18 (50.0) | < 0.001 | 21 (43.8) | 8 (16.7) | 0.008 |
| PG [mg/dL] | 96.0 [92.0–105.0] | 94.0 [89.0–99.0] | 0.004 | 96.8 ± 8.2 | 92.1 ± 5.7 | 0.007 | 101.0 [94.0–109.2] | 95.0 [92.8–102.2] | 0.026 |
| HbA1c [%] | 5.6 [5.4–5.8] | 5.5 [5.3–5.6] | 0.002 | 5.7 ± 0.3 | 5.6 ± 0.2 | 0.114 | 6.0 [5.8–6.1] | 5.7 [5.5–5.9] | < 0.001 |
| HG, n (%) | 36 (37.1) | 2 (2.1) | < 0.001 | 15 (41.7) | 0 (0.0) | < 0.001 | 37 (77.1) | 6 (12.5) | < 0.001 |
| Diabetes, n (%) | 2 (2.1) | 0 (0.0) | 0.497 | 0 (0.0) | 0 (0.0) | — | 7 (14.6) | 3 (6.2) | 0.316 |
| MetS component, n (%) | — | — | < 0.001 | — | — | < 0.001 | — | — | < 0.001 |
| 0 | 0 (0.0) | 18 (18.6) | — | 0 (0.0) | 16 (44.4) | — | 0 (0.0) | 19 (39.6) | — |
| 1 | 0 (0.0) | 74 (76.3) | — | 0 (0.0) | 19 (52.8) | — | 0 (0.0) | 26 (54.2) | — |
| 2 | 88 (90.7) | 5 (5.2) | — | 33 (91.7) | 1 (2.8) | — | 42 (87.5) | 3 (6.2) | — |
| 3 | 9 (9.3) | 0 (0.0) | — | 3 (8.3) | 0 (0.0) | — | 6 (12.5) | 0 (0.0) | — |
| TyG-WC | 812.3 ± 63.8 | 728.6 ± 75.9 | < 0.001 | 845.3 ± 71.5 | 768.2 ± 84.1 | < 0.001 | 798.6 ± 52.3 | 743.4 ± 59.5 | < 0.001 |
| TyG-BMI | 227.5 ± 23.6 | 203.2 ± 25.6 | < 0.001 | 226.2 [218.3–235.8] | 209.9 [192.8–235.4] | 0.008 | 222.5 ± 18.9 | 204.8 ± 21.7 | < 0.001 |

Values are presented as mean ± SD or median [interquartile range], depending on data distribution. Categorical variables are expressed as number (%). “MetS component, n (%)” indicates the number of components meeting the diagnostic criteria for MetS among the following: HT, DLP, HG, excluding OB. Abbreviations: MetS, metabolic syndrome; WC, waist circumference; BMI, body mass index; SBP, systolic blood pressure; DBP, diastolic blood pressure; HT, hypertension; TG, triglycerides; HDL-C, high-density lipoprotein cholesterol; DLP, dyslipidemia; PG, plasma glucose; HbA1c, glycated hemoglobin; HG, hyperglycemia; TyG-WC, triglyceride-glucose index × waist circumference; TyG-BMI, triglyceride-glucose index × body mass index.

**S10 Table. Clinical characteristics of MetS and control groups at year 0 in Clusters 4–6.**

| | Cluster4 MetS | Cluster4 Control | Cluster4 p-value | Cluster5 MetS | Cluster5 Control | Cluster5 p-value | Cluster6 MetS | Cluster6 Control | Cluster6 p-value |
|---|---|---|---|---|---|---|---|---|---|
| n | 44 | 44 | — | 46 | 46 | — | 25 | 25 | — |
| WC [cm] | 86.2 [85.6–86.9] | 81.8 [79.8–84.0] | < 0.001 | 86.5 [85.5–87.7] | 81.8 [79.8–84.8] | < 0.001 | 86.4 [85.6–87.5] | 80.9 [79.3–81.8] | < 0.001 |
| WC ≥ 85 cm, n (%) | 44 (100.0) | 6 (13.6) | < 0.001 | 46 (100.0) | 10 (21.7) | < 0.001 | 25 (100.0) | 1 (4.0) | < 0.001 |
| BMI [kg/m$^2$] | 23.4 [22.8–24.3] | 22.8 [21.2–23.5] | 0.002 | 23.8 ± 1.5 | 22.6 ± 1.8 | < 0.001 | 23.8 ± 1.8 | 21.2 ± 1.5 | < 0.001 |
| BMI ≥ 25, n (%) | 2 (4.5) | 2 (4.5) | 1.0 | 10 (21.7) | 4 (8.7) | 0.147 | 6 (24.0) | 0 (0.0) | 0.022 |
| SBP [mmHg] | 134.7 ± 14.6 | 132.0 ± 14.8 | 0.390 | 137.0 [130.2–143.8] | 131.0 [122.0–136.0] | 0.006 | 127.0 [121.0–133.0] | 129.0 [124.0–136.0] | 0.627 |
| DBP [mmHg] | 86.7 ± 11.2 | 84.0 ± 1.0 | 0.231 | 86.9 ± 7.9 | 83.6 ± 9.2 | 0.067 | 81.2 ± 5.9 | 82.6 ± 9.8 | 0.556 |
| HT, n (%) | 41 (93.2) | 28 (63.6) | 0.002 | 45 (97.8) | 31 (67.4) | < 0.001 | 16 (64.0) | 20 (80.0) | 0.345 |
| TG [mg/dL] | 200.0 [136.8–255.5] | 142.5 [112.8–192.5] | 0.008 | 128.5 [92.8–165.0] | 85.5 [62.2–108.5] | < 0.001 | 201.0 [164.0–255.0] | 133.0 [88.0–200.0] | 0.067 |
| HDL-C [mg/dL] | 51.5 [41.0–63.0] | 56.5 [48.0–64.5] | 0.080 | 58.1 ± 13.4 | 64.9 ± 13.4 | 0.018 | 51.0 ± 9.5 | 55.0 ± 10.9 | 0.176 |
| DLP, n (%) | 40 (90.9) | 26 (59.1) | 0.001 | 24 (52.2) | 5 (10.9) | < 0.001 | 23 (92.0) | 19 (76.0) | 0.247 |
| PG [mg/dL] | 96.7 ± 9.4 | 92.9 ± 9.5 | 0.065 | 102.4 ± 10.7 | 99.5 ± 12.2 | 0.230 | 110.0 [100.0–124.0] | 105.0 [99.0–120.0] | 0.522 |
| HbA1c [%] | 5.7 ± 0.2 | 5.6 ± 0.3 | 0.052 | 5.8 [5.7–6.0] | 5.6 [5.5–5.8] | 0.002 | 6.2 [6.1–6.5] | 5.8 [5.6–6.2] | 0.003 |
| HG, n (%) | 11 (25.0) | 5 (11.4) | 0.167 | 27 (58.7) | 7 (15.2) | < 0.001 | 25 (100.0) | 15 (60.0) | 0.001 |
| Diabetes, n (%) | 0 (0.0) | 1 (2.3) | 1.000 | 2 (4.3) | 3 (6.5) | 1.000 | 12 (48.0) | 9 (36.0) | 0.567 |
| MetS component, n (%) | — | — | < 0.001 | — | — | < 0.001 | — | — | 0.051 |
| 0 | 0 (0.0) | 5 (11.4) | — | 0 (0.0) | 10 (21.7) | — | 0 (0.0) | 0 (0.0) | — |
| 1 | 0 (0.0) | 20 (45.5) | — | 0 (0.0) | 30 (65.2) | — | 0 (0.0) | 4 (16.0) | — |
| 2 | 40 (90.9) | 18 (40.9) | — | 42 (91.3) | 5 (10.9) | — | 11 (44.0) | 13 (52.0) | — |
| 3 | 4 (9.1) | 1 (2.3) | — | 4 (8.7) | 1 (2.2) | — | 14 (56.0) | 8 (32.0) | — |
| TyG-WC | 786.3 ± 50.2 | 717.2 ± 50.7 | < 0.001 | 759.5 ± 41.4 | 686.0 ± 55.3 | < 0.001 | 809.6 ± 70.8 | 723.1 ± 72.5 | < 0.001 |
| TyG-BMI | 213.3 ± 15.5 | 196.0 ± 19.7 | < 0.001 | 207.6 ± 16.7 | 188.5 ± 20.3 | < 0.001 | 221.8 ± 26.0 | 190.5 ± 19.4 | < 0.001 |

Values are presented as mean ± SD or median [interquartile range], depending on data distribution. Categorical variables are expressed as number (%). “MetS component, n (%)” indicates the number of components meeting the diagnostic criteria for MetS among the following: HT, DLP, HG, excluding OB. Abbreviations: MetS, metabolic syndrome; WC, waist circumference; BMI, body mass index; SBP, systolic blood pressure; DBP, diastolic blood pressure; HT, hypertension; TG, triglycerides; HDL-C, high-density lipoprotein cholesterol; DLP, dyslipidemia; PG, plasma glucose; HbA1c, glycated hemoglobin; HG, hyperglycemia; TyG-WC, triglyceride-glucose index × waist circumference; TyG-BMI, triglyceride-glucose index × body mass index.

**S11 Table. Lifestyle characteristics of MetS and control groups in Clusters 1–3 (year −3 to −1).**

| | Cluster1 MetS | Cluster1 Control | Cluster1 p-value | Cluster2 MetS | Cluster2 Control | Cluster2 p-value | Cluster3 MetS | Cluster3 Control | Cluster3 p-value |
|---|---|---|---|---|---|---|---|---|---|
| n | 97 | 97 | — | 36 | 36 | — | 48 | 48 | — |
| Frequency of smoking | 0.0 [0.0–1.0] | 0.0 [0.0–0.7] | 0.188 | 0.7 [0.0–1.0] | 0.0 [0.0–1.0] | 0.052 | 0.0 [0.0–1.0] | 0.2 [0.0–1.0] | 0.371 |
| Smoking cessation, n (%) | 2 (2.1) | 2 (2.1) | 1.0 | 3 (8.3) | 0 (0.0) | 0.239 | 1 (2.1) | 4 (8.3) | 0.362 |
| Frequency of daily drinking | 1.0 [0.0–1.0] | 1.0 [0.0–1.0] | 0.818 | 0.0 [0.0–1.0] | 0.0 [0.0–1.0] | 0.614 | 0.0 [0.0–1.0] | 0.0 [0.0–1.0] | 0.646 |
| Frequency of lifestyle efforts | 0.0 [0.0–0.3] | 0.0 [0.0–0.3] | 0.709 | 0.0 [0.0–0.2] | 0.0 [0.0–0.7] | 0.082 | 0.0 [0.0–0.3] | 0.0 [0.0–0.3] | 0.909 |
| Willingness to receive guidance, n (%) | 32 (34.0) | 45 (46.9) | 0.078 | 15 (41.7) | 13 (37.1) | 0.809 | 23 (52.3) | 22 (48.9) | 0.833 |

All continuous variables were non-normally distributed and are presented as median [interquartile range]. Categorical variables are expressed as number (%). For each variable, participants with missing values were excluded from the variable-specific denominators. The group-specific sample sizes shown in the row labeled “n” represent the total number of participants in each group, including those with missing lifestyle data.

**S12 Table. Lifestyle characteristics of MetS and control groups in Clusters 1–3 (year −1 to 0).**

| | Cluster1 MetS | Cluster1 Control | Cluster1 p-value | Cluster2 MetS | Cluster2 Control | Cluster2 p-value | Cluster3 MetS | Cluster3 Control | Cluster3 p-value |
|---|---|---|---|---|---|---|---|---|---|
| n | 97 | 97 | — | 36 | 36 | — | 48 | 48 | — |
| Smoking cessation, n (%) | 5 (5.2) | 0 (0.0) | 0.059 | 1 (2.8) | 0 (0.0) | 1.000 | 2 (4.2) | 0 (0.0) | 0.495 |
| Shift toward daily drinking, n (%) | 4 (4.2) | 3 (3.1) | 0.721 | 1 (2.8) | 1 (2.8) | 1.0 | 0 (0.0) | 1 (2.2) | 1.000 |
| Ceased daily drinking, n (%) | 2 (2.1) | 3 (3.1) | 1.000 | 0 (0.0) | 1 (2.8) | 1.000 | 1 (2.2) | 2 (4.3) | 1.000 |
| Reduced lifestyle effort, n (%) | 9 (9.9) | 7 (7.4) | 0.608 | 5 (13.9) | 0 (0.0) | 0.054 | 2 (4.4) | 4 (8.7) | 0.677 |
| Willingness to receive guidance, n (%) | 35 (37.6) | 49 (51.6) | 0.058 | 16 (44.4) | 15 (42.9) | 1.000 | 25 (56.8) | 23 (50.0) | 0.534 |

Categorical variables are expressed as number (%).

For each variable, participants with missing values were excluded from the variable-specific denominators. The group-specific sample sizes shown in the row labeled “n” represent the total number of participants in each group, including those with missing lifestyle data.

**S13 Table. Lifestyle characteristics of MetS and control groups in Clusters 4–6 (year −3 to −1).**

| | Cluster4 MetS | Cluster4 Control | Cluster4 p-value | Cluster5 MetS | Cluster5 Control | Cluster5 p-value | Cluster6 MetS | Cluster6 Control | Cluster6 p-value |
|---|---|---|---|---|---|---|---|---|---|
| n | 44 | 44 | — | 46 | 46 | — | 25 | 25 | — |
| Frequency of smoking | 0.0 [0.0–1.0] | 0.0 [0.0–1.0] | 0.379 | 0.7 [0.0–1.0] | 0.0 [0.0–1.0] | 0.247 | 0.0 [0.0–1.0] | 0.0 [0.0–0.0] | 0.105 |
| Smoking cessation, n (%) | 3 (6.8) | 1 (2.3) | 0.616 | 0 (0.0) | 0 (0.0) | — | 0 (0.0) | 0 (0.0) | — |
| Frequency of daily drinking | 0.0 [0.0–1.0] | 1.0 [0.3–1.0] | 0.016 | 1.0 [0.0–1.0] | 1.0 [0.0–1.0] | 0.846 | 0.3 [0.0–1.0] | 0.0 [0.0–1.0] | 0.689 |
| Frequency of lifestyle efforts | 0.0 [0.0–0.6] | 0.0 [0.0–0.3] | 0.688 | 0.0 [0.0–0.3] | 0.0 [0.0–0.3] | 0.466 | 0.0 [0.0–0.3] | 0.0 [0.0–0.3] | 0.772 |
| Willingness to receive guidance, n (%) | 21 (50.0) | 20 (50.0) | 1.000 | 16 (37.2) | 23 (54.8) | 0.130 | 8 (34.8) | 8 (32.0) | 1.0 |

All continuous variables were non-normally distributed and are presented as median [interquartile range]. Categorical variables are expressed as number (%). For each variable, participants with missing values were excluded from the variable-specific denominators. The group-specific sample sizes shown in the row labeled “n” represent the total number of participants in each group, including those with missing lifestyle data.

**S14 Table. Lifestyle characteristics of MetS and control groups in Clusters 4–6 (year −1 to 0).**

| | Cluster4 MetS | Cluster4 Control | Cluster4 p-value | Cluster5 MetS | Cluster5 Control | Cluster5 p-value | Cluster6 MetS | Cluster6 Control | Cluster6 p-value |
|---|---|---|---|---|---|---|---|---|---|
| n | 44 | 44 | — | 46 | 46 | — | 25 | 25 | — |
| Smoking cessation, n (%) | 3 (6.8) | 1 (2.3) | 0.616 | 5 (10.9) | 2 (4.3) | 0.434 | 3 (12.5) | 0 (0.0) | 0.110 |
| Shift toward daily drinking, n (%) | 3 (7.0) | 0 (0.0) | 0.242 | 0 (0.0) | 4 (9.5) | 0.050 | 1 (4.2) | 2 (8.0) | 1.000 |
| Ceased daily drinking, n (%) | 2 (4.7) | 0 (0.0) | 0.495 | 1 (2.2) | 0 (0.0) | 1.000 | 0 (0.0) | 1 (4.0) | 1.000 |
| Reduced lifestyle effort, n (%) | 8 (18.6) | 5 (12.5) | 0.551 | 2 (4.4) | 3 (7.1) | 0.669 | 4 (17.4) | 1 (4.0) | 0.180 |
| Willingness to receive guidance, n (%) | 21 (50.0) | 20 (50.0) | 1.000 | 20 (45.5) | 25 (58.1) | 0.286 | 8 (34.8) | 9 (36.0) | 1.000 |

Categorical variables are expressed as number (%).

For each variable, participants with missing values were excluded from the variable-specific denominators. The group-specific sample sizes shown in the row labeled “n” represent the total number of participants in each group, including those with missing lifestyle data.

| | Year −3 | | | | Year −2 | | | | Year −1 | | | |
|---|---|---|---|---|---|---|---|---|---|---|---|---|
| Pearson | WC | HT | DLP | HG | WC | HT | DLP | HG | WC | HT | DLP | HG |
| A | 0 | 1 | 1 | 0 | 0 | 2 | 1 | 0 | 0 | 2 | 2 | 0 |
| B | 0 | 1 | 0 | 0 | 1 | 1 | 0 | 0 | 2 | 1 | 0 | 0 |
| C | 2 | 1 | 1 | 0 | 2 | 2 | 1 | 0 | 2 | 2 | 0 | 0 |

Vectorization

| Pearson | Vector |
|---|---|
| A | [0,1,1,0,0,2,1,0,0,2,2,0] |
| B | [0,1,0,0,1,1,0,0,2,1,0,0] |
| C | [2,1,1,0,2,2,1,0,2,2,0,0] |

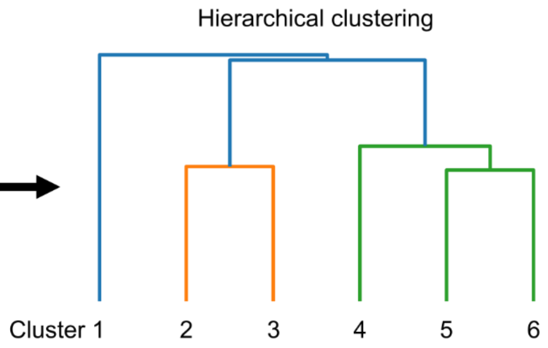


**S1 Fig. Encoding process of MetS components for clustering analysis.**

Abbreviations: MetS, metabolic syndrome; WC, waist circumference (three-level classification); HT, hypertension (three-level classification); DLP, dyslipidemia (three-level classification); HG, hyperglycemia (three-level classification).

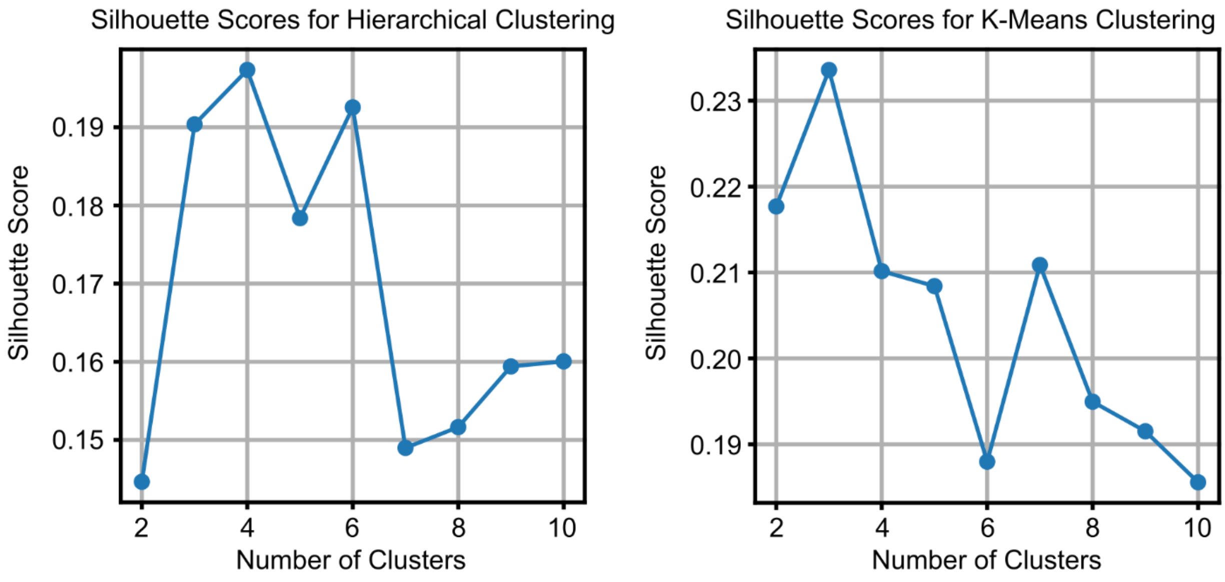


Cluster by K-Means Clustering (columns); Cluster by Hierarchical Clustering (rows)

| | 1 | 2 | 3 | 4 | 5 | 6 |
|---|---|---|---|---|---|---|
| 1 | 11 | 2 | 79 | 0 | 5 | 0 |
| 2 | 3 | 0 | 0 | 32 | 0 | 1 |
| 3 | 5 | 0 | 4 | 2 | 0 | 37 |
| 4 | 0 | 42 | 0 | 2 | 0 | 0 |
| 5 | 2 | 4 | 4 | 0 | 31 | 5 |
| 6 | 0 | 13 | 0 | 8 | 2 | 2 |

ARI = 0.597

Cluster by K-Means Clustering (columns); Cluster by Hierarchical Clustering (rows)

| | 1 | 2 | 3 | 4 | 5 | 6 | 7 |
|---|---|---|---|---|---|---|---|
| 1 | 9 | 0 | 75 | 0 | 6 | 0 | 7 |
| 2 | 2 | 0 | 0 | 33 | 0 | 0 | 1 |
| 3 | 5 | 0 | 4 | 2 | 0 | 37 | 0 |
| 4 | 0 | 20 | 0 | 2 | 0 | 0 | 22 |
| 5 | 2 | 16 | 4 | 0 | 19 | 5 | 0 |
| 6 | 0 | 17 | 0 | 7 | 0 | 1 | 0 |

ARI = 0.522

**S2 Fig. Silhouette scores and ARI for cluster number evaluation and method comparison.**

Abbreviations: ARI, adjusted Rand index.

**MetS**

| | Cluster 1 | | | | Cluster 2 | | | | Cluster 3 | | | |
|---|---|---|---|---|---|---|---|---|---|---|---|---|
| Year | -3 | -2 | -1 | 0 | -3 | -2 | -1 | 0 | -3 | -2 | -1 | 0 |
| None | 1.0% | 3.1% | 1.0% | – | 2.8% | 2.8% | – | – | – | 2.1% | – | – |
| HT | 15.5% | 16.5% | 13.4% | – | 2.8% | – | – | – | – | – | – | – |
| DLP | 1.0% | – | – | – | 11.1% | 19.4% | – | – | – | – | – | – |
| HG | – | – | – | – | – | – | – | – | 2.1% | 2.1% | 2.1% | – |
| HT, DLP | 1.0% | 3.1% | 1.0% | – | – | – | – | – | – | – | – | – |
| HT, HG | – | – | – | – | – | – | – | – | – | – | – | – |
| DLP, HG | – | – | – | – | – | – | – | – | – | – | – | – |
| HT, DLP, HG | – | – | – | – | – | – | – | – | – | – | – | – |
| OB | 7.2% | 9.3% | 3.1% | – | 22.2% | 2.8% | 8.3% | – | 47.9% | 47.9% | 52.1% | – |
| OB, HT | 72.2% | 67.0% | 81.4% | – | – | – | – | – | 14.6% | 14.6% | 14.6% | – |
| OB, DLP | 2.1% | 1.0% | – | – | 58.3% | 75.0% | 86.1% | – | 12.5% | 4.2% | 4.2% | – |
| OB, HG | – | – | – | – | 2.8% | – | 5.6% | – | 22.9% | 29.2% | 27.1% | – |
| OB, HT, DLP | – | – | – | 62.9% | – | – | – | 58.3% | – | – | – | 22.9% |
| OB, HT, HG | – | – | – | 26.8% | – | – | – | 2.8% | – | – | – | 56.2% |
| OB, DLP, HG | – | – | – | 1.0% | – | – | – | 30.6% | – | – | – | 8.3% |
| OB, HT, DLP, HG | – | – | – | 9.3% | – | – | – | 8.3% | – | – | – | 12.5% |

| | Cluster 4 | | | | Cluster 5 | | | | Cluster 6 | | | |
|---|---|---|---|---|---|---|---|---|---|---|---|---|
| Year | -3 | -2 | -1 | 0 | -3 | -2 | -1 | 0 | -3 | -2 | -1 | 0 |
| None | 2.3% | 2.3% | 2.3% | – | 34.8% | 21.7% | 17.4% | – | – | 16.0% | – | – |
| HT | 6.8% | 4.5% | 6.8% | – | 45.7% | 39.1% | 34.8% | – | – | – | – | – |
| DLP | 36.4% | 11.4% | 9.1% | – | – | – | – | – | 20.0% | 8.0% | 12.0% | – |
| HG | – | – | – | – | 6.5% | 6.5% | 6.5% | – | 12.0% | 4.0% | 8.0% | – |
| HT, DLP | 50.0% | 68.2% | 68.2% | – | – | 6.5% | 4.3% | – | 4.0% | – | – | – |
| HT, HG | 2.3% | 2.3% | 2.3% | – | 6.5% | 10.9% | 13.0% | – | 20.0% | 16.0% | 16.0% | – |
| DLP, HG | – | – | 2.3% | – | – | 2.2% | – | – | 16.0% | 24.0% | 28.0% | – |
| HT, DLP, HG | – | 2.3% | 6.8% | – | – | 2.2% | 2.2% | – | 28.0% | 32.0% | 32.0% | – |
| OB | – | – | – | – | – | 4.3% | 4.3% | – | – | – | – | – |
| OB, HT | – | 4.5% | – | – | 6.5% | 6.5% | 10.9% | – | – | – | – | – |
| OB, DLP | 2.3% | 4.5% | 2.3% | – | – | – | 4.3% | – | – | – | 4.0% | – |
| OB, HG | – | – | – | – | – | – | 2.2% | – | – | – | – | – |
| OB, HT, DLP | – | – | – | 75.0% | – | – | – | 41.3% | – | – | – | – |
| OB, HT, HG | – | – | – | 9.1% | – | – | – | 47.8% | – | – | – | 8.0% |
| OB, DLP, HG | – | – | – | 6.8% | – | – | – | 2.2% | – | – | – | 36.0% |
| OB, HT, DLP, HG | – | – | – | 9.1% | – | – | – | 8.7% | – | – | – | 56.0% |

**S3 Fig. Heatmaps showing the distribution of MetS component combinations across Clusters 1–6 in the MetS group.** Clusters 1–3 (top) and Clusters 4–6 (bottom) are displayed vertically. Each row represents a unique combination of MetS diagnostic components, including OB, HT, DLP, and HG. “None” indicates individuals who met none of the diagnostic criteria for any of the four components. Each column corresponds to a specific year within each cluster, from year −3 to 0 (left to right). Cell values indicate the proportion of individuals with each component combination, calculated as a percentage of the total number of individuals in that column. The percentage is annotated within each cell. Color intensity reflects the proportion, ranging from 0% (lightest) to 100% (darkest). Cells with no individuals are marked with an en dash (–).

Abbreviations: MetS, metabolic syndrome; OB, abdominal obesity; HT, hypertension; DLP, dyslipidemia; HG, hyperglycemia.

**Control**

| | Cluster 1 | | | | Cluster 2 | | | | Cluster 3 | | | |
|---|---|---|---|---|---|---|---|---|---|---|---|---|
| Year | -3 | -2 | -1 | 0 | -3 | -2 | -1 | 0 | -3 | -2 | -1 | 0 |
| None | 1.0% | 7.2% | 6.2% | 7.2% | 2.8% | 16.7% | 16.7% | 22.2% | – | 10.4% | 10.4% | 10.4% |
| HT | 15.5% | 18.6% | 24.7% | 24.7% | 2.8% | 2.8% | 2.8% | – | – | 6.2% | 12.5% | 6.2% |
| DLP | 1.0% | 1.0% | – | 2.1% | 11.1% | 13.9% | 16.7% | 13.9% | – | – | – | 2.1% |
| HG | – | – | – | – | – | – | – | – | 4.2% | 2.1% | 2.1% | – |
| HT, DLP | 1.0% | 3.1% | 5.2% | 3.1% | – | – | 2.8% | 2.8% | – | – | – | – |
| HT, HG | – | – | – | 2.1% | – | – | – | – | – | – | – | 4.2% |
| DLP, HG | – | – | – | – | – | – | – | – | – | – | 2.1% | 2.1% |
| HT, DLP, HG | – | – | – | – | – | – | – | – | – | – | – | – |
| OB | 7.2% | 9.3% | 11.3% | 11.3% | 27.8% | 25.0% | 30.6% | 22.2% | 47.9% | 31.2% | 22.9% | 29.2% |
| OB, HT | 72.2% | 55.7% | 48.5% | 47.4% | – | 2.8% | 5.6% | 5.6% | 14.6% | 20.8% | 20.8% | 27.1% |
| OB, DLP | 2.1% | 5.2% | 3.1% | 2.1% | 52.8% | 33.3% | 22.2% | 33.3% | 12.5% | 18.8% | 14.6% | 12.5% |
| OB, HG | – | – | 1.0% | – | 2.8% | 5.6% | 2.8% | – | 20.8% | 10.4% | 14.6% | 6.2% |
| OB, HT, DLP | – | – | – | – | – | – | – | – | – | – | – | – |
| OB, HT, HG | – | – | – | – | – | – | – | – | – | – | – | – |
| OB, DLP, HG | – | – | – | – | – | – | – | – | – | – | – | – |
| OB, HT, DLP, HG | – | – | – | – | – | – | – | – | – | – | – | – |

| | Cluster 4 | | | | Cluster 5 | | | | Cluster 6 | | | |
|---|---|---|---|---|---|---|---|---|---|---|---|---|
| Year | -3 | -2 | -1 | 0 | -3 | -2 | -1 | 0 | -3 | -2 | -1 | 0 |
| None | 2.3% | 18.2% | 20.5% | 9.1% | 34.8% | 21.7% | 28.3% | 17.4% | – | 8.0% | 8.0% | – |
| HT | 6.8% | 15.9% | 15.9% | 18.2% | 45.7% | 43.5% | 30.4% | 39.1% | – | 12.0% | 12.0% | – |
| DLP | 36.4% | 15.9% | 9.1% | 15.9% | – | 2.2% | – | 4.3% | 20.0% | 12.0% | 12.0% | 8.0% |
| HG | – | – | 2.3% | – | 6.5% | 6.5% | 6.5% | 4.3% | 12.0% | – | – | 4.0% |
| HT, DLP | 50.0% | 38.6% | 36.4% | 34.1% | – | – | – | 4.3% | 4.0% | 12.0% | 16.0% | 28.0% |
| HT, HG | 2.3% | – | 2.3% | 4.5% | 6.5% | 10.9% | 8.7% | 6.5% | 20.0% | 24.0% | 8.0% | 20.0% |
| DLP, HG | – | 2.3% | – | 2.3% | – | – | – | – | 16.0% | 12.0% | 12.0% | 4.0% |
| HT, DLP, HG | – | – | 2.3% | 2.3% | – | – | 4.3% | 2.2% | 28.0% | 16.0% | 24.0% | 32.0% |
| OB | – | – | – | 2.3% | – | 6.5% | 4.3% | 4.3% | – | – | – | – |
| OB, HT | – | 2.3% | 6.8% | 4.5% | 6.5% | 6.5% | 15.2% | 15.2% | – | – | – | – |
| OB, DLP | 2.3% | 6.8% | 4.5% | 4.5% | – | 2.2% | – | – | – | – | – | 4.0% |
| OB, HG | – | – | – | 2.3% | – | – | 2.2% | 2.2% | – | 4.0% | 8.0% | – |
| OB, HT, DLP | – | – | – | – | – | – | – | – | – | – | – | – |
| OB, HT, HG | – | – | – | – | – | – | – | – | – | – | – | – |
| OB, DLP, HG | – | – | – | – | – | – | – | – | – | – | – | – |
| OB, HT, DLP, HG | – | – | – | – | – | – | – | – | – | – | – | – |

**S4 Fig. Heatmaps showing the distribution of MetS component combinations across Clusters 1–6 in the control group.** Clusters 1–3 (top) and Clusters 4–6 (bottom) are displayed vertically. Each row represents a unique combination of MetS diagnostic components, including OB, HT, DLP, and HG. “None” indicates individuals who met none of the diagnostic criteria for any of the four components. Each column corresponds to a specific year within each cluster, from year −3 to 0 (left to right). Cell values indicate the proportion of individuals with each component combination, calculated as a percentage of the total number of individuals in that column. The percentage is annotated within each cell. Color intensity reflects the proportion, ranging from 0% (lightest) to 100% (darkest). Cells with no individuals are marked with an en dash (–).

Abbreviations: OB, abdominal obesity; HT, hypertension; DLP, dyslipidemia; HG, hyperglycemia.

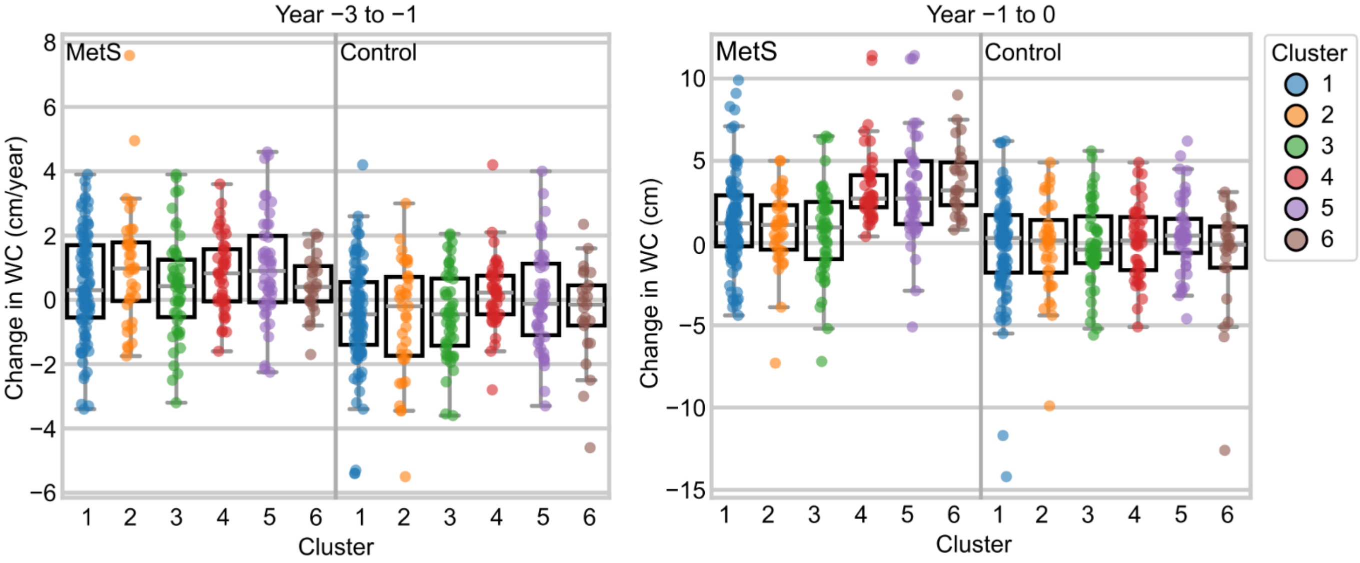


**S5 Fig. Cluster-specific WC change.**

Abbreviations: MetS, metabolic syndrome; WC, waist circumference.

**MetS and control groups**

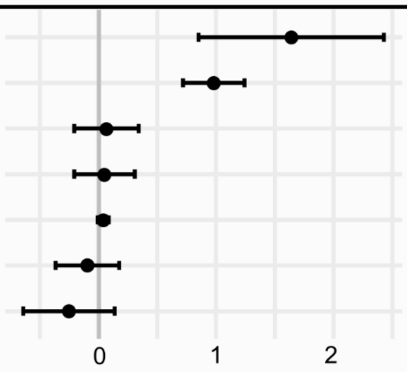


| Variables | Beta | SE | 95% CI | P-value |
|---|---|---|---|---|
| Smoking cessation | 1.64 | 0.40 | [0.85–2.43] | <0.001 |
| MetS | 0.98 | 0.13 | [0.72–1.24] | <0.001 |
| Frequency of smoking | 0.07 | 0.14 | [−0.21–0.34] | 0.634 |
| Willingness to receive guidance | 0.05 | 0.13 | [−0.21–0.31] | 0.710 |
| Age | 0.04 | 0.02 | [−0.01–0.09] | 0.103 |
| Frequency of daily drinking | −0.10 | 0.14 | [−0.37–0.18] | 0.492 |
| Frequency of lifestyle efforts | −0.25 | 0.20 | [−0.64–0.14] | 0.203 |

**Adjusted $R^2$ = 0.112**

**MetS group**

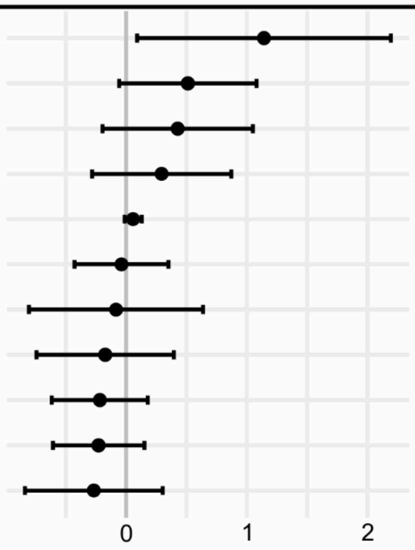


| Variables | Beta | SE | 95% CI | P-value |
|---|---|---|---|---|
| Smoking cessation | 1.14 | 0.54 | [0.09–2.19] | 0.034 |
| Cluster 5 | 0.51 | 0.29 | [−0.06–1.08] | 0.079 |
| Cluster 2 | 0.43 | 0.32 | [−0.19–1.05] | 0.179 |
| Cluster 4 | 0.30 | 0.29 | [−0.28–0.87] | 0.317 |
| Age | 0.06 | 0.04 | [−0.01–0.13] | 0.105 |
| Frequency of smoking | −0.04 | 0.20 | [−0.43–0.35] | 0.848 |
| Cluster 6 | −0.08 | 0.37 | [−0.80–0.64] | 0.822 |
| Cluster 3 | −0.17 | 0.29 | [−0.74–0.40] | 0.553 |
| Frequency of daily drinking | −0.22 | 0.20 | [−0.61–0.18] | 0.286 |
| Willingness to receive guidance | −0.23 | 0.19 | [−0.61–0.15] | 0.240 |
| Frequency of lifestyle efforts | −0.27 | 0.29 | [−0.84–0.30] | 0.361 |

**Adjusted $R^2$ = 0.026**

**S6 Fig. Predictors of annual WC change from year −3 to −1.** Results are based on multiple linear regression analyses examining factors associated with annual changes in WC. The top panel shows results for the combined group (MetS and control), and the bottom panel shows results for the MetS group alone. Definitions and descriptions of the lifestyle variables are provided in S1–S3 Tables.

Abbreviations: MetS, metabolic syndrome; WC, waist circumference.

**MetS and control groups**

| Variables | Beta | SE | 95% CI | P-value |
|---|---|---|---|---|
| Smoking cessation | 2.36 | 0.64 | [1.11–3.62] | <0.001 |
| MetS | 1.86 | 0.25 | [1.37–2.35] | <0.001 |
| Shift toward daily drinking | 0.56 | 0.65 | [−0.72–1.84] | 0.391 |
| Reduced lifestyle effort | 0.28 | 0.43 | [−0.56–1.12] | 0.517 |
| Age | −0.08 | 0.04 | [−0.16–0.01] | 0.092 |
| Willingness to receive guidance | −0.41 | 0.24 | [−0.89–0.06] | 0.088 |
| Ceased daily drinking | −1.39 | 0.80 | [−2.95–0.18] | 0.084 |

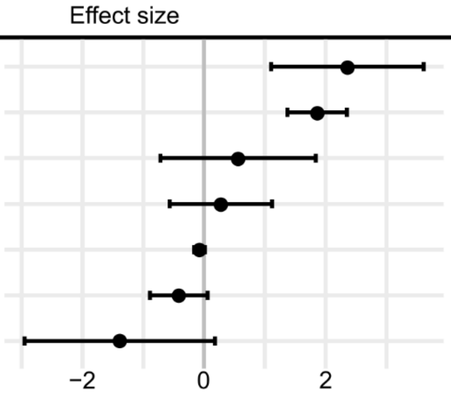


**Adjusted $R^2$ = 0.146**

**MetS group**

| Variables | Beta | SE | 95% CI | P-value |
|---|---|---|---|---|
| Cluster 6 | 2.31 | 0.64 | [1.06–3.56] | <0.001 |
| Cluster 4 | 2.07 | 0.51 | [1.08–3.07] | <0.001 |
| Smoking cessation | 1.75 | 0.65 | [0.48–3.02] | 0.007 |
| Cluster 5 | 1.75 | 0.50 | [0.77–2.74] | 0.001 |
| Ceased daily drinking | 1.47 | 1.13 | [−0.74–3.68] | 0.194 |
| Shift toward daily drinking | 0.27 | 0.93 | [−1.56–2.10] | 0.773 |
| Reduced lifestyle effort | 0.03 | 0.54 | [−1.03–1.10] | 0.950 |
| Age | −0.04 | 0.06 | [−0.16–0.09] | 0.559 |
| Cluster 2 | −0.46 | 0.54 | [−1.51–0.59] | 0.393 |
| Willingness to receive guidance | −0.53 | 0.33 | [−1.18–0.12] | 0.114 |
| Cluster 3 | −0.55 | 0.50 | [−1.54–0.43] | 0.273 |

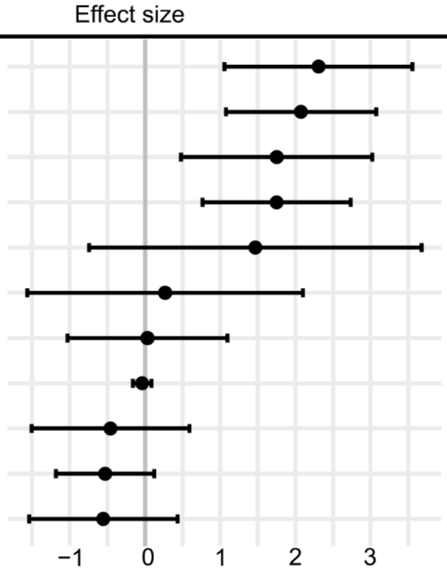


**Adjusted $R^2$ = 0.161**

**S7 Fig. Predictors of annual WC change from year −1 to 0.** Results are based on multiple linear regression analyses examining factors associated with annual changes in WC. The top panel shows results for the combined group (MetS and control), and the bottom panel shows results for the MetS group alone. Definitions and descriptions of the lifestyle variables are provided in S1–S3 Tables.

Abbreviations: MetS, metabolic syndrome; WC, waist circumference.

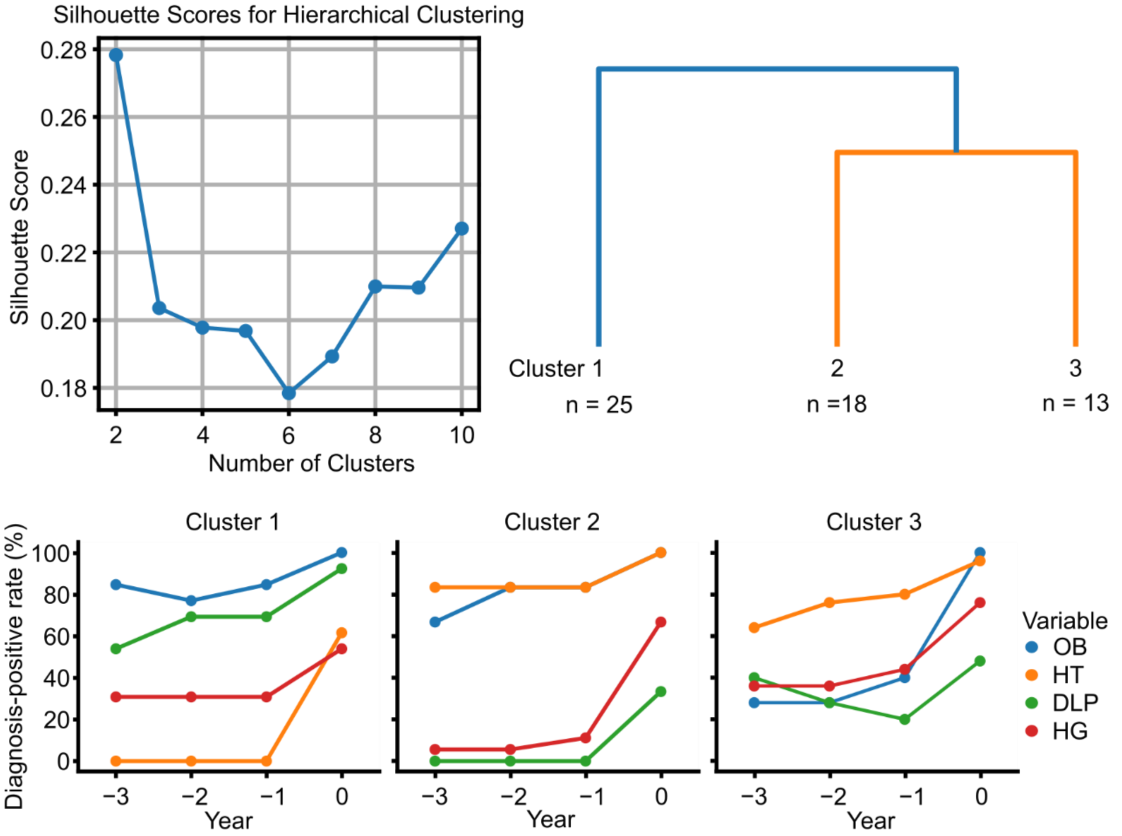


**S8 Fig. Diagnosis-positive rates for each MetS component—OB, HT, DLP, and HG—in females.**

Females included in the analysis were n = 56 (median age: 55.0 years; interquartile range: 54.0–57.0), with age values corresponding to year −3.

Abbreviations: MetS, metabolic syndrome; OB, abdominal obesity; HT, hypertension; DLP, dyslipidemia; HG, hyperglycemia.

Abbreviations: MetS, metabolic syndrome; OB, abdominal obesity; HT, hypertension; DLP, dyslipidemia; HG, hyperglycemia.

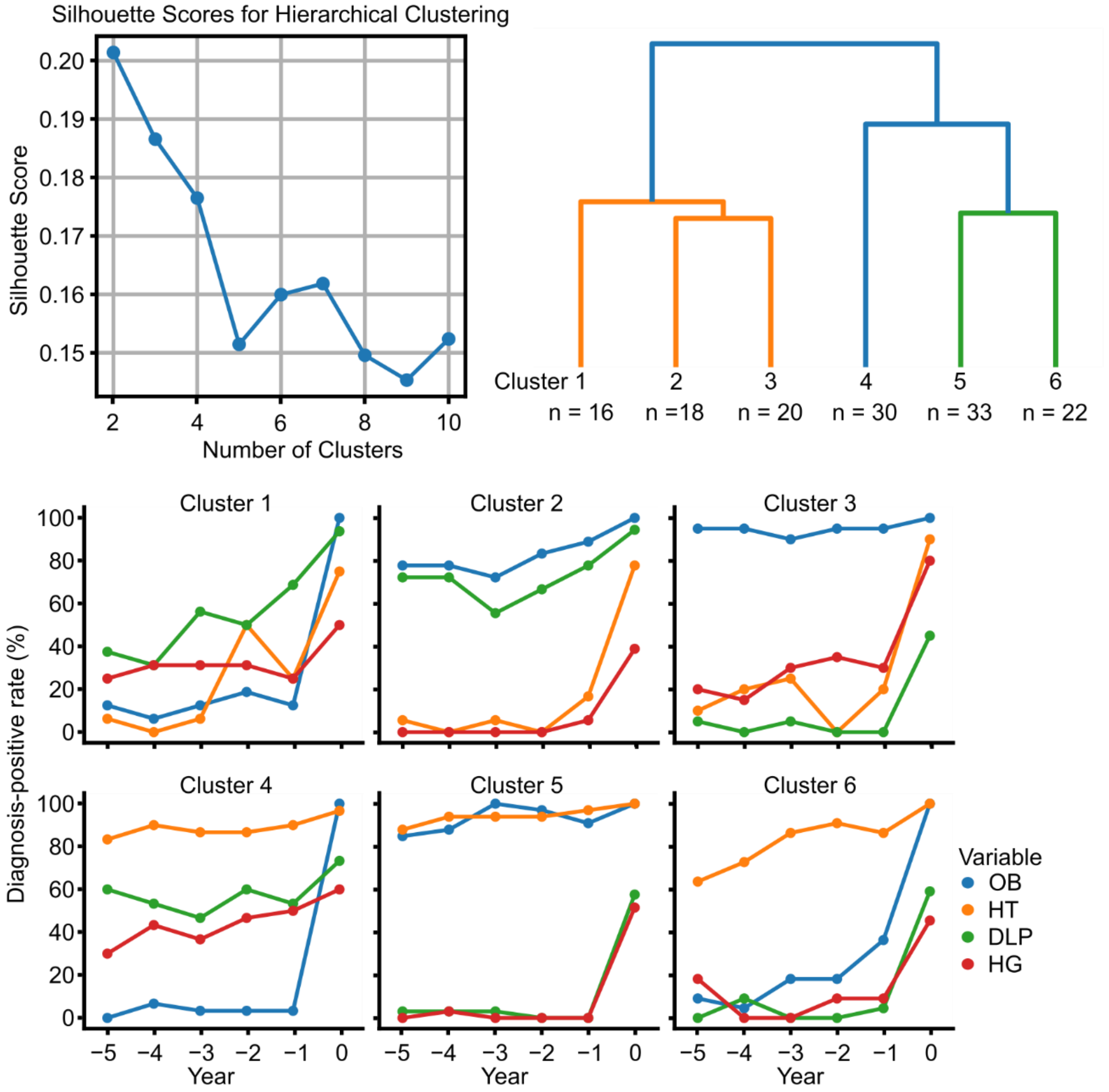


**S9 Fig. Diagnosis-positive rates for each MetS component—OB, HT, DLP, and HG—in males, with analyses covering the period from year −5 to 0.**

Males included in the analysis were n = 139 (median age: 54.0 years; interquartile range: 52.0–55.0), with age values corresponding to year −5.

Abbreviations: MetS, metabolic syndrome; OB, abdominal obesity; HT, hypertension; DLP, dyslipidemia; HG, hyperglycemia.

Abbreviations: MetS, metabolic syndrome; OB, abdominal obesity; HT, hypertension; DLP, dyslipidemia; HG, hyperglycemia.
